# Multi-trait genome-wide association study in 34,394 Chinese women reveals the genetic architecture of plasma metabolites during pregnancy

**DOI:** 10.1101/2023.11.26.23299022

**Authors:** Siyang Liu, Jilong Yao, Liang Lin, Xianmei Lan, Linlin Wu, Nannan Kong, Yuqing Deng, Yan Li, Jiansheng Xie, Huanhuan Zhu, Xiaoxia Wu, Zilong Li, Likuan Xiong, Yuan Wang, Jinghui Ren, Xuemei Qiu, Weihua Zhao, Ya Gao, Yuanqing Chen, Fengxia Su, Yun Zhou, Weiqiao Rao, Jing Zhang, Guixue Hou, Liping Huang, Linxuan Li, Xinhong Liu, Chao Nie, Liqiong Luo, Zengyou Liu, Fang Chen, Shengmou Lin, Lijian Zhao, Qingmei Fu, Dan Jiang, Ye Yin, Xun Xu, Jian Wang, Huanming Yang, Rong Wang, Jianmin Niu, Fengxiang Wei, Xin Jin, Siqi Liu

## Abstract

Metabolites are important indicators of individual health and can serve as crucial targets for therapy. However, the genetic basis of many metabolites remains largely unexplored, especially among underrepresented East Asians and during critical periods such as pregnancy. In this study, we utilized genetic information obtained from non-invasive prenatal testing to conduct a genome-wide association analysis of 84 metabolites, including 37 amino acids, 10 vitamins, 24 metal elements, and 13 hormones, among 34,394 Chinese pregnant women. Of these metabolites, 52 and 11 had not previously been studied in East Asians or globally. We identified 30 novel metabolite-gene associations. We also observed substantial differences in the genetic effects on hormones between pregnancy and non-pregnancy periods, suggesting effect modifications in response to physiological changes. Furthermore, we uncovered pervasive pleiotropic effects for 50.94% of the genetic associations among metabolites, as well as between six metabolites and eight pregnancy biomarkers. Using mendelian randomization, we identified potential causal relationships between plasma folate and ischemic stroke, vitamin D3 and Graves’ disease, copper and open-angle glaucoma, and androstenedione and rheumatoid arthritis. These discoveries provide invaluable genetic insights into human metabolism, laying the foundation for future mechanistic studies and the development of new therapeutic targets, particularly for underrepresented East Asians.

## Introduction

Metabolites are small molecules that act as intermediate or end products during the metabolic processes^1^. Their concentrations are influenced by both genetic and environmental factors^2,3^. Metabolites can have a crucial role in disease etiology and are often targeted in therapeutic interventions^4,5^. Human genetics provides an important approach to understanding the determinants of metabolite alteration and assessing the role of metabolites in disease outcomes^6^. Genome-wide association studies (GWAS) of metabolites have identified thousands of variants associated with approximately 7,000 metabolites in blood plasma or urine, as reported in the latest GWAS catalog (r2023-01-30)^7^. However, like many other traits, human genetic studies of metabolites have been dominated by European populations^8^. Specifically, less than 300 metabolites (less than 4.29% of all metabolites) have been studied in East Asian populations, with generally small sample sizes (less than 2,000 individuals)^9–16^. Therefore, there is a significant gap in our understanding of the genetic effects of metabolites in underrepresented East Asian populations.

Previous genetic studies of metabolites have not yet addressed the question of whether and how genetic effects may vary during pregnancy, a special and critical period that plays a pivotal role in human reproduction. Pregnancy involves a series of physiological changes and metabolic adaptations that necessitate a balanced metabolome for the health of both the mother and the fetus^17^. Observational epidemiological studies have demonstrated that inadequate or excessive metabolite concentrations can result in poor health outcomes or severe disease and adverse birth outcomes. For example, excessive vitamin A levels may cause fetal malformation^18^, a lack of folic acid (vitamin B9) was associated with a high risk of neural tube defects^19^, a deficiency of selenium was related to an elevated risk of miscarriage, pre-eclampsia, and growth restriction^20,21^, and vitamin D insufficiency was related to fetal bone growth retardation ^22^. To investigate the causal relationship and molecular mechanisms underlying these observational associations, genetic studies of plasma metabolite levels during pregnancy are crucial. However, maternal metabolites during pregnancy have not been explored using genome-wide association studies, and the genetic basis of metabolite levels during pregnancy is largely unknown. It is pertinent to investigate whether the genetic effects on metabolites, as estimated from genome-wide association studies, may differ between pregnant and non-pregnant populations. Moreover, it is important to investigate whether metabolites assayed during this critical early period of human life may have a causal relationship with diseases that manifest later in life. Such investigations will yield critical insights into human metabolism, providing the foundation for downstream mechanistic studies and the identification of new therapeutic targets.

Conducting large-scale genetic analyses, particularly in underrepresented populations, remains a significant challenge. In a prior pioneering study, we developed methodologies and protocols for robust genetic association analysis using non-invasive prenatal testing (NIPT) data.^23^. In this study, we conducted genetic analyses of a set of 84 maternal metabolites (abbreviated as MM) during pregnancy, leveraging genetic resources obtained from NIPT and metabolite measurements from tandem mass spectrometry among 34,394 Chinese pregnant women. These 84 metabolites included 37 amino acids, 5 hydrophilic vitamins, 5 hydrophobic vitamins, 13 hormones, and 24 trace elements and heavy metals. Of these metabolites, 52 have not been studied in East Asians, and 11 have not been investigated globally before (Table S1). The exact number of participants involved for each metabolite was detailed in Table S2. We performed both single- and multi-trait genome-wide association analyses to identify genetic signals that significantly contribute to plasma metabolite levels and to estimate SNP heritability. We compared the genetic effects of the same metabolite measurements estimated from both pregnancy participants and non-pregnancy individuals. Additionally, we conducted wald ratio estimation and colocalization analyses to identify pleiotropic effects among metabolites in the context of the KEGG pathway and between metabolites and several pregnancy phenotypes reported in a companion study. Finally, we applied mendelian randomization to investigate the potential causal effects of pregnancy metabolites on disease traits in East Asian populations. This study represents the largest scale metabolite genome-wide association study in underrepresented East Asian populations and has provided the first insight into the genetic basis of the plasma level of 84 metabolites during pregnancy.

## Results

### Study design and phenotypic distribution of the 84 metabolites

The 34, 394 pregnant participants were recruited during their routine obstetric examinations in the city of Shenzhen, South China. The expense of the non-invasive prenatal test (NIPT) is covered by the government and the insurance in the Shenzhen city. When NIPT is provided, each participant was asked if they would like to partake in the pregnancy nutrition program for screening of metabolites. We did not identify a a difference of ages between participants partaking in the nutrition programs and those did not. The mean age of the participants was 30 years (standard deviation [s.d.] 4.9), and the mean gestational week for blood sampling was 16 weeks plus 3 days (s.d. 4.1 weeks) (Table S2). Non-invasive prenatal tests (NIPT) were provided as a routine prenatal screening test^24^, while tandem mass spectrometry (MS/MS) assays of 84 metabolites covering four categories were offered as an additional screening option to monitor the metabolite status of the participants using the same tube of blood plasma.

For the NIPT test, whole genome low-pass sequencing (0.1x - 0.3x) was carried out, and we applied NIPT sequencing of the 34,394 participants using two sequencing platforms to minimize potential technical artifacts due to low-pass sequencing. Specifically, 18,091 of them were sequenced using the blackbird sequencing machine (BB) and 16,303 were sequenced using the Seq500 sequencing machine (Seq500). We performed variation calling, genotype imputation, and genome-wide association tests for data from each platform before we move on to the meta-analysis and the multi-trait association study. The genotype imputation was performed using an adaptive hidden Markov model with a Chinese population reference panel, and we achieved imputation accuracy of 0.758 and 0.892 for BB and Seq500 data, respectively (see Methods).

In the MS/MS assay, due to limited plasma volume, we separated the samples for different assays including amino acids, hormones, water-soluble vitamins (WSV), fat-soluble vitamins (FSV), and metal elements. The plasma volume limited our capability to examine each metabolite for all the 34,394 participants. The mean effective sample size was 7595, 10973, 8225 and 12033 for amino acid, hormone, vitamin and metal elements, respectively and was detailed in Table S2. The plasma concentration of 84 metabolites followed a Gaussian distribution with the presence of outliers in a few of the metal elements (**Figure 1**). The mean value of the metabolites fell in the reference ranges except for 3MHis, Al, I, Ti, ALD, and vitamin D, which may be due to specific physiological status during pregnancy (Table S2).

**Figure 1.**
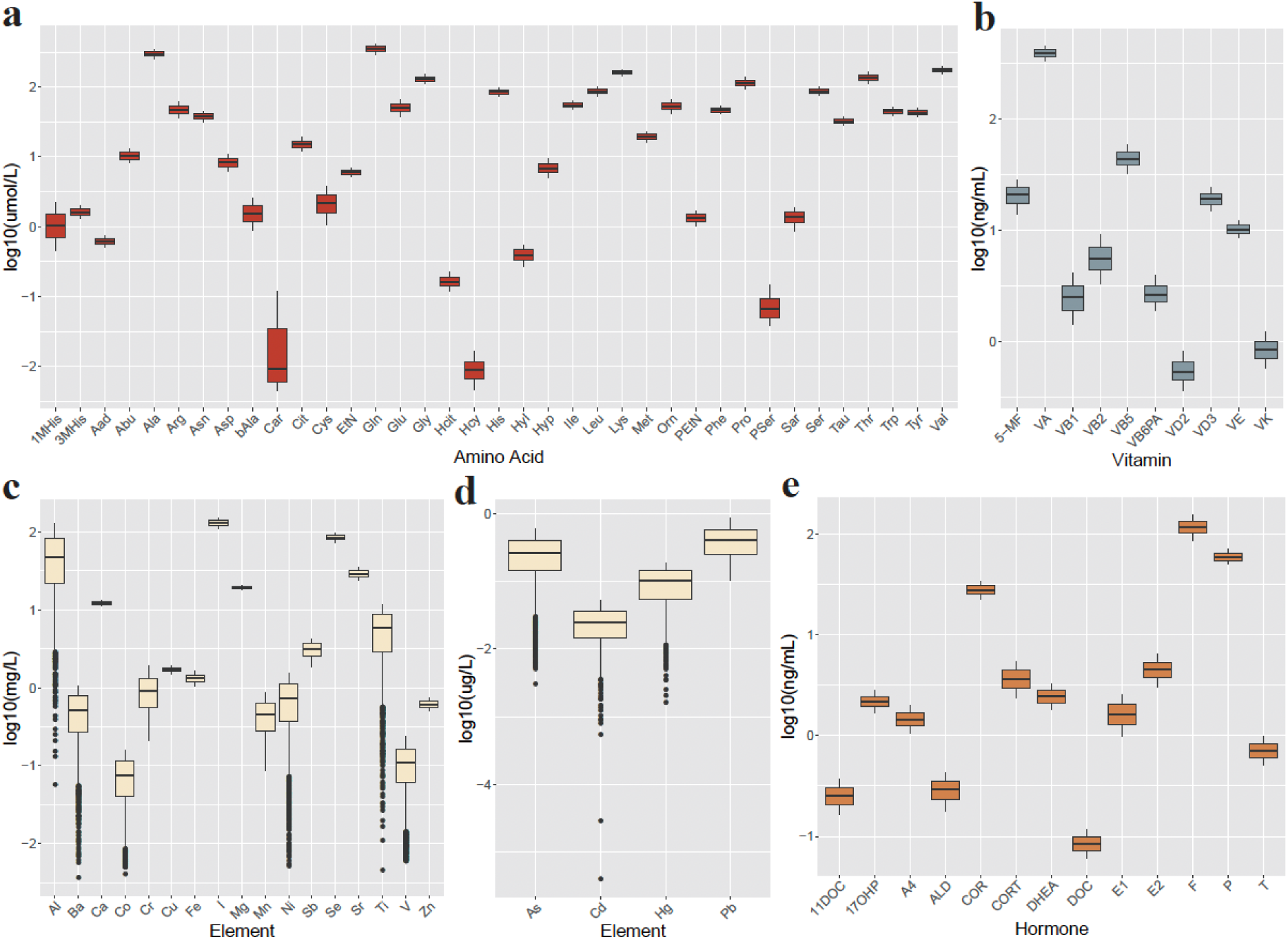
Phenotypic distribution of the 84 metabolites. Shown were the logarithm of the concentration for a) 37 amino acids b) 10 vitamins c) and d) 24 elements and e) 13 hormones. Units of the metabolites were shown in the title of the y-axis. The outliners were excluded in the genome-wide association analysis.

The metabolites tend to function in a pathway, and therefore we suspected that metabolites that play a role together in a specific pathway would be correlated with each other. The phenotypic correlation between the metabolites suggests that metabolites that belong to the same category tend to be more phenotypically correlated with each other than metabolites that belong to different categories (Figure S1, Table S3). Strong phenotypic correlations were observed between Met and His, among Phe, Ile/Leu, and Tyr/Val for the amino acids, between Cd and Hg, among Al, Ga, and V for metal elements, and between A4 and T, and between T1 and T2 for hormones. The phenotypic correlation was smaller for metabolites between different categories, with the strongest correlation observed between VA and PSer/Sar (Spearman’s R ∼ 0.41) and between VE and Car (Spearman’s R ∼ 0.42).

### Genome-wide association analysis identifies 53 metabolite-gene associations

Based on the abovementioned observation of the phenotypic correlation within the same metabolite categories, we conducted a multi-trait genome-wide association study for each metabolite by utilizing information obtained from GWAS of metabolites from the same category. Firstly, we performed a regression analysis using PLINK^25^, to regress the quantile-transformed metabolite value on the genotype dosage for BB and Seq500 sequencing data, respectively. The analysis was adjusted for confounding factors, including the first three principal components, maternal age, gestational week of non-invasive prenatal testing (NIPT), MS/MS testing, and the inferred sex of the fetus. Detailed information can be found in Table S4. Subsequently, we conducted a meta-analysis using METAL^26^ to combine GWAS summary statistics from BB and Seq500. Finally, we utilized the multi-trait association test algorithm implemented in MTAG^27^ estimates for each SNP, using information from summary statistics of metabolites within the same metabolite categories.

The genomic control lambda (GC lambda) for the 84 metabolite molecules ranged from 0.9 to 1.05, indicating a negligible inflation of association statistics. In total, we identified 53 genetic associations reaching genome-wide significance (p<5x10^-8^) for 33 metabolites, including 14 amino acids, 7 elements, 6 hormones, and 7 vitamins (**Figure 2**). Among the 53 loci, 23 were previously known to be associated with the corresponding metabolite level, while 30 were novel discoveries. The largest standardized effect for each of the four metabolite categories was observed at (1) the *CPS1-ERBB4* locus (lead SNP rs75472842, β_nor_=-0.46, where an additional copy of the T allele decreases 60.92 μmol/L glycine in maternal plasma); (2) the *CXorf47* locus (lead SNP rs1804495, β_nor_=-0.34, where an additional copy the A allele decreases 43.75 μg/L iodine); (3) the *ZNF468 locus* (lead SNP rs4801940, β_nor_=-0.20, where an additional copy of the T allele decreases 0.45 ng/mL 17OHP); and (4) the *LINC00441 locus* ( rs144131853, β_nor_=-0.32, where an additional copy of the T allele decreases 0.32 ng/mL of vitamin D3) (**Table 1**). The complete information for the 53 genetic loci can be found in Table S5. We present the Manhattan, quantile-quantile, and locuszoom plots for the 53 metabolite-gene associations in Figure S2. Below, we summarize the key findings for each metabolite category.

**Figure 2.**
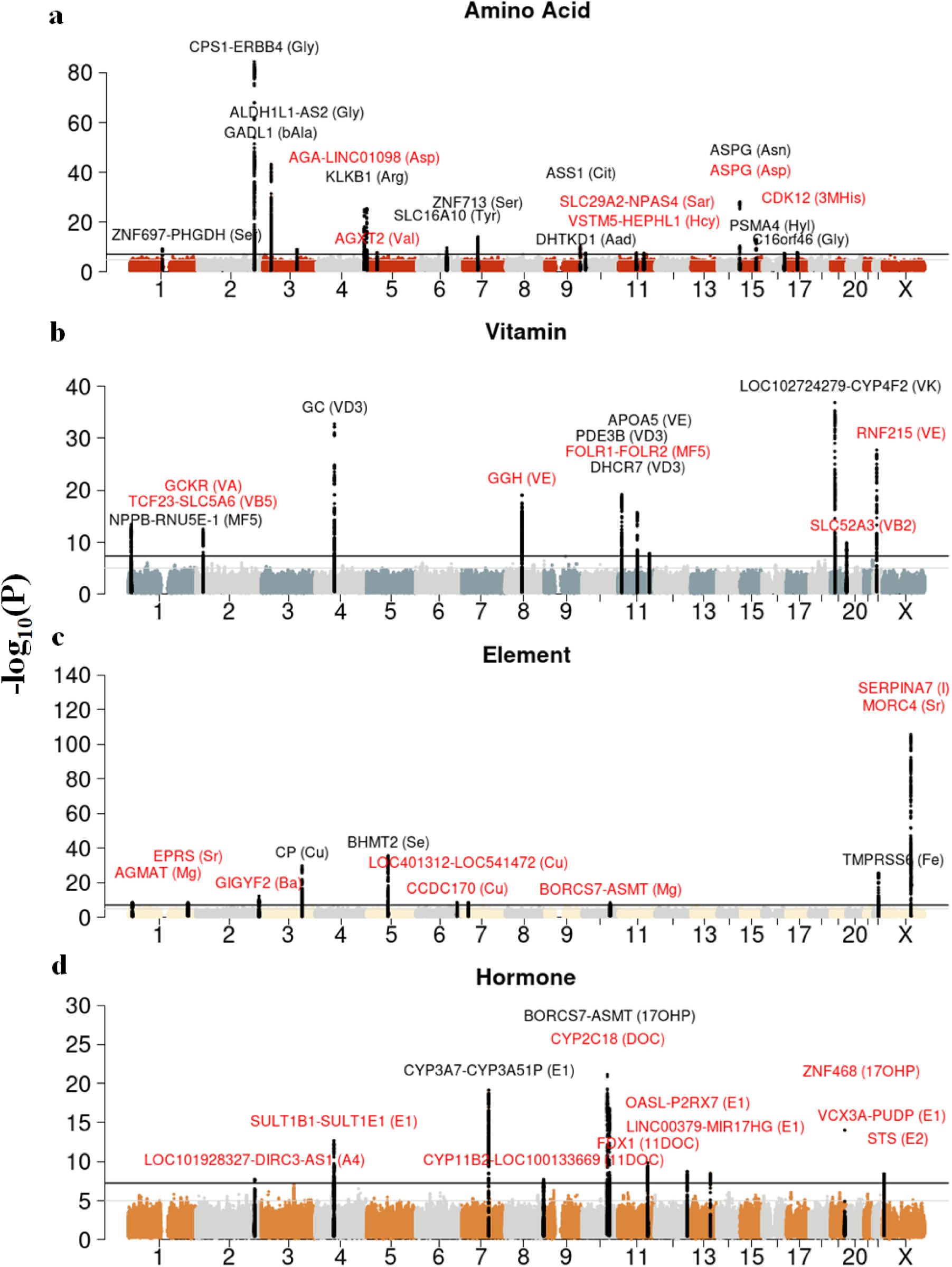
The 53 significant genetic associations with the metabolites. Manhattan plot of all the genetic associations for amino acids (a), vitamins (b), metal elements (c) and hormones (d). The x-axis is the chromosomal position and the y-axis represents the minus logarithmic transformation of the *P* value from the GWAS regression model. The black and grey horizontal line represents the *P* value of the genome-wide significance level at *P*=5.0×10^-8^ and *P*=1.0×10^-5^. Each significant locus is annotated by the nearest gene symbol with the corresponding trait in parenthesis. Gene symbols in red suggest that the genetic association locus (1Mbp centering on the lead SNP) was not previously identified in the GWAS catalog or PubMed. Gene symbols in black reflect known associations.

**Table 1.**
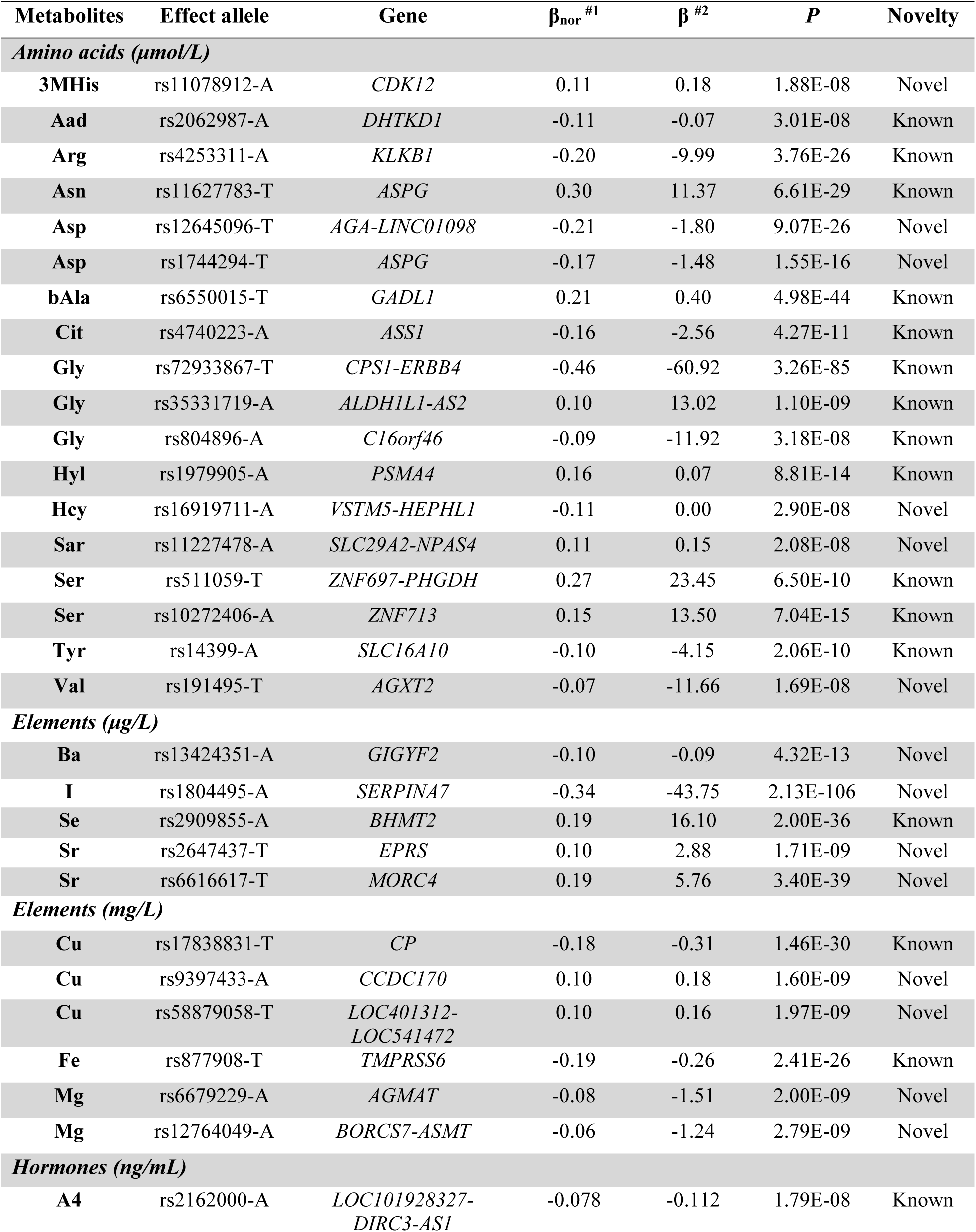
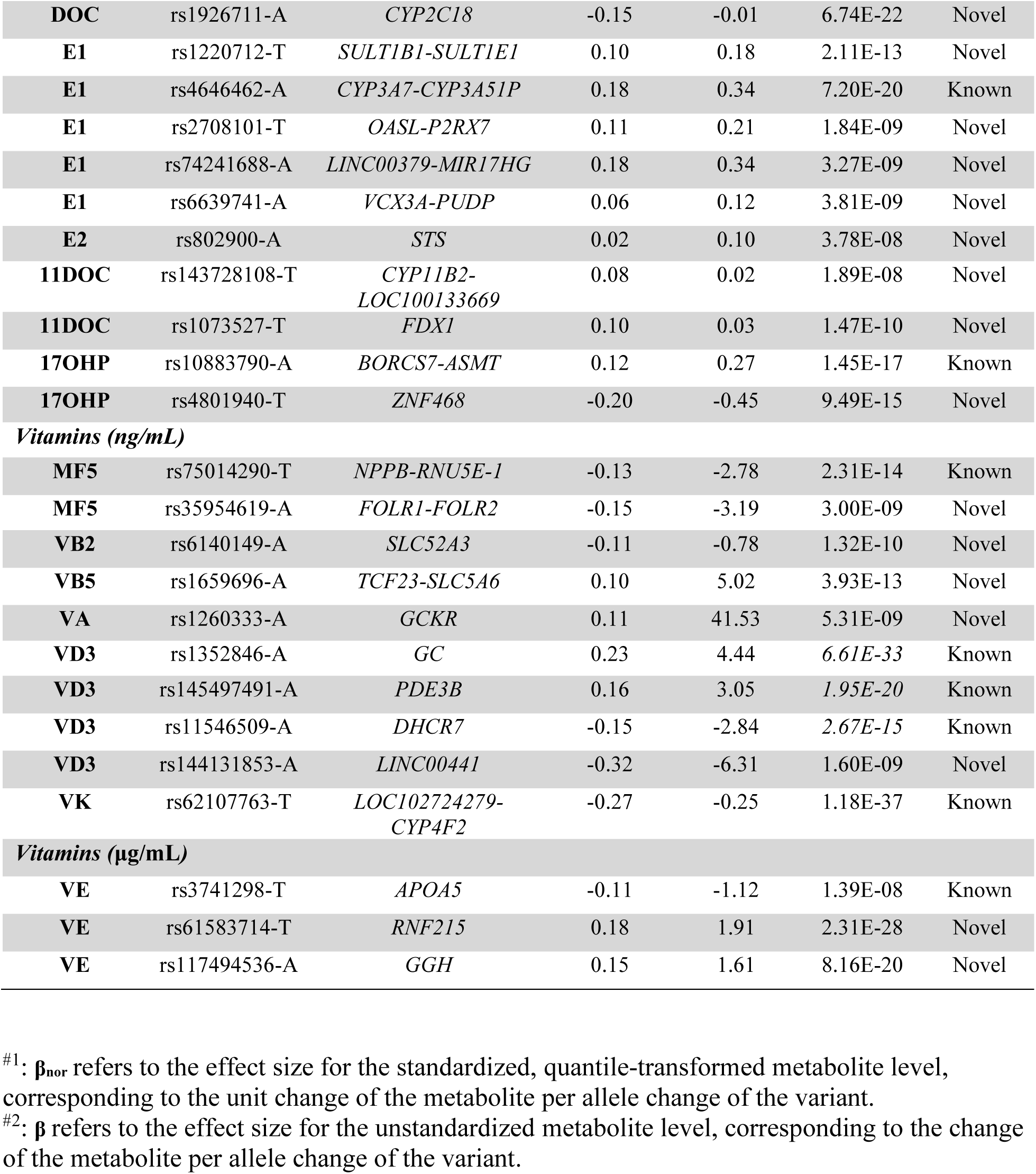
The fifty-three genetic loci associated with maternal plasma metabolites.

### Amino acids

We found 18 independent association signals for 14 amino acids (**Figure 2a**, **Table 1**). All signals were first discovered in the East Asian population, and 12 were previously reported in the European population and demonstrated consistent effect estimates according to the GWAS catalog or PhenoScanner (Table S5). We focused on the interpretation of the six novel association signals found for six amino acids. The first novel signal was found for the 3-Methylhistidine (3MHis), an amino acid biomarker for muscle protein turnover at the *CDK12* locus (lead SNP rs11078912-A: beta=0.07, 95% CI: 0.04 to 0.09, P= 1.88×10^-8^) (**Figure 2a**, Table S5). The rs11078912 variant was an expression quantitative trait locus (eQTL) for *FBL20*, which plays a role in protein metabolism^28^ and was strongly associated with HDL, eGFR, and diseases such as asthma and rheumatoid arthritis^29^. However, this association had not been previously reported, likely due to limited genetic studies (only four small-scale GWAS studies were documented for 3MHis in the GWAS catalog).

Two novel association signals were identified between the *AGA* locus (lead SNP rs12645096-T: beta=-0.13, 95% CI: -0.11 to -0.16, P= 9.07×10^-26^), the *ASPG* locus (lead SNP rs1744294-T: beta=-0.13, 95% CI: -0.10 to -0.17, P= 1.54×10^-16^), and aspartic acid (Asp). rs12645096 is a strong eQTL for *AGA*, which encodes the aspartylglucosaminidase in many tissues such as artery, thyroid, and lungs (p-value < 1.7e-32) in GTEx. Similarly, rs1744294 is also an eQTL for *ASPG* that encodes asparaginase. Both aspartylglucosaminidase and asparaginase deficiencies can lead to an increased Asn and Asp, which explained the genetic association observed in our study. Those associations had not been previously reported due to limited genetic studies on this metabolite^29,30^. While Asp has been used for the treatment of fatigue and improvement of athletic performance and muscle strength without good scientific evidence supporting its use^31^, the identified genetic association may serve as instrumental variables for causal inference of its utility when combined with data from biobanks with genomic data.

The fourth novel genetic association was identified for homocysteine (Hcy) at the intergenic *VSTM5*-*HEPHL1* locus (lead SNP rs16919711-A: beta=-0.08, 95% CI: - 0.12 to -0.05, P= 2.90×10^-8^). The rs16919711 variant was an eQTL for *PANX1* and *GPR83* in cultured fibroblasts cells and the cerebellar hemisphere in the brain. However, the summary statistics of the previous 14 GWAS on Hcy were not available for download, and we were not able to evaluate the effect size of rs16919711 in other studies. As Hcy is an important biomarker for cardiovascular and cerebrovascular disease^32^, it will be worthwhile to replicate and validate the function of this novel association signal.

Finally, we found a novel association between sarcosine (Sar) and the intergenic *SLC29A2*-*NPAS4* locus (lead SNP rs11227478-A: beta=0.08, 95% CI: 0.05 to 0.11, P= 2.08×10^-8^) and an association between Valine (Val) and the *AGXT2* locus (lead SNP rs191495-T: beta=-0.05, 95% CI: -0.03 to -0.07, P= 1.69×10^-8^). rs11227478 is an eQTL for *CTSF* in several tissues and has been reported to associate with height and forced vital capacity^29^. There was limited knowledge of rs191495 at the mitochondrial aminotransferase *AGXT2.* Information from future functional studies is critical for interpreting these observed genetic associations.

### Vitamins

We identified 12 loci that have a strong impact on the levels of seven vitamins in maternal plasma, six of which are known associations and six of which are novel associations (**Figure 1b**). We have confirmed the previously known association between the 5-methyltetrahydrofolate level (MF5) and the *NPPB-RNU5E-1* locus (lead SNP rs75014290, P= 9.49×10^-15^), which is located near *MTHFR*, the gene that encodes the methylenetetrahydrofolate reductase. We also identified a new association between MF5 and variants within the *FOLR1-FOLR2* gene locus (lead SNP rs35954619-A: beta=-0.09, 95% CI: -0.06 to -0.12, P= 3.00×10^-9^), an eQTL for *FOLR1* which encodes the folate receptor 1 and folate receptor beta, and was previously associated with the asymmetrical dimethylarginine levels related to heart rate variability ^33^. On average, individuals with an additional T and A alleles for rs75014290 and rs35954619, respectively, exhibit 2.78 ng/mL and 3.19 ng/ml lower 5-methyltetrahydrofolate in maternal plasma during pregnancy (see **Table 1**).

We have also identified a new association between vitamin B2 (VB2) and the *SLC52A3* gene (lead SNP rs6140149-A: beta=-0.08, 95% CI: -0.05 to -0.10, P= 1.32×10^-10^), which is known to play a role in water-soluble vitamin metabolism ^34^, as well as a new association between vitamin B5 (VB5) and the *TCF23*-*SLC5A6* locus (lead SNP rs1659696-A: beta=0.10, 95% CI: 0.07 to 0.13, P= 3.92×10^-13^), an eQTL for *ATRAID*, which plays a role in cell cycle arrest in all-trans-retinoic acid signal pathway^28^. The discoveries of the three new genetic associations with the three water-soluble vitamins, despite their strong biological relevance, were due to a lack of previous genome-wide association studies (Table S1).

As for the fat-soluble vitamins, we identified the known associations between 25-Hydroxyvitamin D3 level (VD3) and three loci *GC* (rs1352846, P= 6.61×10^-33^) *PDE3B* (rs10766189, P= 1.95×10^-20^), and *DHCR7* (rs12789751, P= 2.67×10^-15^), as well as two known associations for vitamin E (VE) at the *APOA5* (P= 1.39×10^-8^) and vitamin K (VK) at the *LOC102724279*-*CYP4F2* locus (P= 1.18×10^-37^). Additionally, we have identified a new association for vitamin A (VA) at the *GCKR* locus (lead SNP rs1260333-A: beta=0.07, 95% CI: 0.05 to 0.10, P= 5.31×10^-9^); a new association for VD3 at the *LINC00441* locus (lead SNP rs144131853-A: beta=-0.04, 95% CI: 0.05 to 0.09, P=1.60×10^-9^); and two new associations for VE at the *RNF215* (lead SNP rs61583714-T: beta=0.14, 95% CI: 0.12 to 0.17, P= 2.31×10^-28^) and *GGH*(lead SNP rs117494536-A: beta=0.12, 95% CI: 0.10 to 0.15, P= 8.16×10^-20^) locus. The *LINC00441*, *RNF215,* and the *GGH* loci were eQTL for *RB1* in adipose tissue, *SEC14L3* in Thyroid, and *GGH* in the whole blood, suggesting biological relevance.

### Metal elements

We identified eleven genetic associations for seven different metal elements, including three previously reported and eight novel signals (**Figure 2b**). The three known loci were the *CP* locus for copper level (Cu) (lead SNP rs17838831: P= 1.46×10^-30^), the *TMPRSS6* locus for iron level (Fe) (lead SNP rs877908: P= 2.41×10^-26^), and the *BHMT2* locus for selenium level (Se) (lead SNP rs2909855: P= 2.00×10^-^ 36). The eight novel associations included the *GIGYF2* locus, an eQTL for *C2orf82* for the barium level (Ba) through lead SNP rs13424351-A (beta=-0.10, 95% CI: -0.07 to -0.12, P= 4.32×10^-13^); the *CCDC170* locus (lead SNP rs9397433-A: beta=-0.07, 95% CI: -0.05 to -0.09, P= 1.60×10^-9^) and the *LOC401312*-*LOC541472* locus, an eQTL for the IL6 antisense RNA 1 (*IL6-AS1*) (lead SNP rs58879058-T: beta=0.07, 95% CI: 0.05 to 0.10, P= 2.00×10^-10^) for copper level (Cu); the *SERPINA7* locus associated with iodine level (I) through lead SNP rs1804495-A, which is a missense variant ( beta=-0.27, 95% CI: -0.25 to -0.30, P= 2.13×10^-106^); the *AGMAT* locus(lead SNP rs6679229-A: beta=-0.06, 95% CI: -0.04 to -0.08, P= 2.00×10^-9^) and the *BORCS7*-*ASMT* locus, an eQTL for *AS3MT* (lead SNP rs12764049-A: beta=-0.06, 95% CI: - 0.04 to -0.08, P= 2.80×10^-9^) were associated with the magnesium level (Mg); and lastly the *EPRS* locus, an eQTL for *EPRS* (lead SNP rs12764049-T: beta=0.07, 95% CI: 0.05 to 0.10, P= 1.71×10^-9^) and the *MORC4* locus, an eQTL for *PRPS1* (lead SNP rs12764049-T: beta=0.16, 95% CI: 0.13 to 0.18, P= 3.40×10^-39^) were associated with strontium level (Sr) in maternal plasma.

The discovery of the strong association between the missense variant rs1804495 at *SERPINA7* and iodine level is noteworthy since no GWAS had been performed previously for iodine. *SERPINA7* encodes the major thyroid hormone transport protein, TBG, in serum. We found that one additional copy of the A allele decreases 11.12 ug/L iodine in maternal plasma, providing evidence for the regulation of iodine metabolism through a common missense genetic polymorphism present in TBG. Lastly, the remaining seven novel genetic associations displayed specific functions and warrant further studies for biological validation.

### Hormone

We identified 12 loci that have a genetic influence on the plasma levels of six hormones, as presented in **Figure 2d**. Those included the known association between the *CYP3A7*-*CYP3A51P* locus and the estrone (E1) (lead SNP rs4646462, P= 7.20×10^-20^) and the association between the *BORCS7*-*ASMT* locus and the 17-hydroxyprogesterone (17OHP) measurement (lead SNP rs10883790, P= 1.45×10^-17^).

We also identified ten novel signals. For instance, the *CYP2C18* locus was found to associate with the 11-Deoxycorticosterone (DOC) (lead SNP rs12764049-A: beta=-0.12, 95% CI: -0.15 to -0.10, P= 6.74×10^-22^). The lead SNP rs1926711 is an eQTL of *CYP2C19*, a monooxygenase that catalyzes many reactions involved in the synthesis of cholesterol, steroids, and other lipids and in drug metabolism.

In addition to the well-known association between the *CYP3A7*-*CYP3A51P* locus and E1, we found four novel signals for E1. These signals included the *SULT1B1*-*SULT1E1* locus (lead SNP rs1220712-T: beta=-0.08, 95% CI: 0.06 to 0.10, P= 2.11×10^-13^), the *OASL*-*P2RX7* locus (lead SNP rs2708101-T: beta=0.07, 95% CI: 0.05 to 0.09, P= 1.84×10^-9^), the *LINC00379-MIR17HG* locus (lead SNP rs74241688-A: beta=0.08, 95% CI: 0.06 to 0.11, P= 3.27×10^-9^) and the *VCX3A*-*PUDP* locus (lead SNP rs6639741-A: beta=0.07, 95% CI: 0.05 to 0.09, P= 3.81×10^-9^).

Furthermore, we discovered a novel signal for Estradiol (E2) levels within *STS* (lead SNP rs802900-: beta=0.06, 95% CI: 0.04 to 0.09, P= 3.78×10^-8^). Similar to the *VCX3A*-*PUDP* locus for E1, this signal is also an eQTL for *PUDP*, which plays a role in nucleotide salvage.

Lastly, we found three novel signals, including two for 11-Deoxycortisol (11DOC) at the *CYP11B2-LOC100133669* (lead SNP rs143728108-T: beta=0.07, 95% CI: 0.04 to 0.09, P= 1.88×10^-8^) and the *FDX1* ( lead SNP rs1073527-T: beta=0.08, 95% CI: 0.06 to 0.10, P= 1.47×10^-10^) locus, which plays roles in the steroid, vitamin D and bile acid metabolism and one signal for 17α-Hydroxyprogesterone (17OHP) at the *ZNF468* locus (lead SNP rs4801940-T: beta=-0.10, 95% CI: -0.08 to -0.13, P= 9.49×10^-15^).

### SNP heritability and genetic correlation

To assess the heritability of the 84 metabolites, we employed the LD score regression (LDSR) technique^35^ to estimate SNP-based heritability. We then compared this estimate to the heritability computed from summary statistics of GWAS from prior investigations^36^ (see Method). Among the 84 metabolites, 67 (78.6%) exhibited a SNP heritability greater than zero (Figure S6, Table S6). The SNP-based heritability estimates ranged from 0.4% to 22.2% with a median of 6.5% the 67 metabolites. Notably, Ala 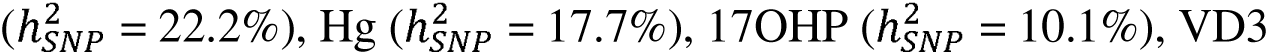 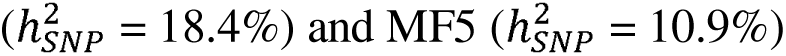 demonstrated the highest SNP heritability among the amino acid, metal elements, hormone, fat, and water vitamin categories, respectively.

When comparing the SNP heritability of 15 metabolites from our study to the publicly available GWAS summary statistics from prior investigations (Kettunen et al., 2016 & Neale’s lab), our study found that all three vitamins, VD3, VA, and VE, as well as the metal element Mg and three amino acids, namely, Tyr, Ile, and Val, exhibited higher SNP heritability (Table S6). However, for the remaining five amino acids and two metal elements, our study found lower SNP heritability (Figure S7).

### Comparison of estimated genetic effects with a non-pregnancy Chinese cohort

All but eight of the 53 genetic loci identified in this study demonstrated the same effect direction and reached nominal significance (*P*<0.05) between the two different sequencing technologies (Black Bird and BGI-Seq500) (**Figure S8,** Table S5). The eight loci that did not reach nominal significance in one of the two sequencing technologies displayed the same effect direction, likely due to lack of power. These results provide support for the robustness of the GWAS hits.

To further explore the possibility of pregnancy-specific genetic associations, we compared the effect size and the p-value for the 53 loci with an independent study of 1,553 non-pregnancy Chinese individuals (Female N=642, Male N=911) from the BGI-Shenzhen multi-omics cohort^37^. These 1,553 participants underwent whole-genome sequencing and a total of 80 out of the 84 metabolites investigated in the present study were also assayed for each individual using the same tandem MS technology(see Methods). Notably in the BGI-Shenzhen multi-omics cohort, amino acids, vitamins, and hormones were assayed using plasma, the same sampling material as this study while the metal elements were assayed using whole blood. A linear regression, adding individual sex and the first two PCs as the covariates, was performed for each of the metabolites (see Method). A regression using only the female individuals was also performed in the non-pregnancy cohort and the results and the conclusions drawn below were consistent.

Interestingly, after excluding four signals for Hcy, Ba, I, and DOC that were not assayed and for Hyl and Se that the lead SNPs were not present in the non-pregnancy cohort, 25 loci discovered in the present study were not replicated in the non-pregnancy cohort (47.2%, *P*>0.05 or effect size were opposite)(Figure S9, Table S7). The inconsistency in genetic effects between the pregnancy and non-pregnancy cohorts varied among the four categories of metabolites. 5 out of the 16 signals in the amino acid category (31.3%), 7 out of the 8 signals in the element category (87.5%), all of the 10 signals in the hormone category (100%), and 2 out of 13 signals in vitamin category (15.4%) demonstrated inconsistent genetic effects between the pregnancy and non-pregnancy participants.

We summarized the reasons for the inconsistency in genetic effects for each category as follows: (1) the low consistency for the element category was likely due to the difference between the plasma and whole blood sampling, as differences in the mean values of the elements between the two populations were observed (Table S7); (2) the low consistency for hormones between the two studies may suggest pregnancy-specific genetic effects, in accordance with the difference in the phenotypic distribution between the two populations or may be due to a lack of power in the non-pregnancy GWAS study; (3) for amino acids, the inconsistency of genetic effect for the *ASPG* locus with Asp, the *SLC29A2*-*NPAS4* locus with Sar, and the *AGXT2* locus with Val were likely due to the differences between the two cohorts, while the inconsistency for the *KLKB1* locus with Arg and the *ASS1* locus with Cit was probably due to lack of power, since the two association signals were previously known. For vitamins, the overall phenotypic distribution between the two populations was similar, except for MF5 and VB5, and therefore the rest of the association signals tended to be consistent between the pregnancy and non-pregnancy cohorts.

### Extensive genetic pleiotropy among metabolites

Metabolites were known to be interacting with each other, playing roles in certain pathways, we inferred the existence of genetic pleiotropy for the metabolites investigated in this study. Specifically, the 84 metabolites examined in this study were involved in 42 KEGG pathways (Table S8) ^38^. The two largest pathways were the aminoacyl-tRNA biosynthesis and steroid hormone biosynthesis pathways, comprising 20 amino acids and 13 hormones in our study. We aim to explore whether the genetic associations we identified suggest pleiotropy.

Among the 53 association signals, 27 (50.94%) affected two or more metabolite (*P* < 0.001, Figure S10). By utilizing a wald ratio estimator to identify the potential causal effect of one metabolite on another using a single SNP^39^, we found 16 genetic associations with a significant effect connecting 18 metabolites (Table S9). The pathway analysis of these 18 metabolites revealed nine pathways with nominal significance (*P* < 0.05) (Table S10). We visualized the pleiotropic effect in the context of the KEGG pathways (**Figure 3**). For instance, in the cyanoamino acid metabolism pathway, the *ASPG* locus (lead SNP rs1744297) suggested an effect from Asp on Asn while the *AGA-LINC01098* locus (lead SNP rs12645096) locus affected both the Asn and Asp. In addition, the *CPS1-ERBB4* locus (lead SNP rs10272406) and *ZNF713* (lead SNP rs72933867) locus suggested an effect from Ser on Gly, different from the direction suggested in the KEGG pathway (**Figure 3a**). In another amino-acid-related aminoacyl-tRNA biosynthesis pathway, *KLKB1* (lead SNP rs4253255) showed an effect from Arg on Met but not the other way around (**Figure 3b**). In the steroid hormone biosynthesis pathway, four gene loci, including *LOC101928327-DIRC3-AS1* (lead SNP rs2162000), *EGR2-NRBF2* (lead SNP rs10822140), *CYP3A5* (lead SNP rs1419745) and *JMJD1C* (lead SNP rs6479891) mutually regulated A4-T-17OHP, E1-E2, E1-E2-CORT, and E1-E2-DHEA, respectively (**Figure 3c**). For the ubiquinone and other terpenoid-quinone biosynthesis pathway that involved vitamins, the *LOC102724279-CYP4F2* locus (lead SNP rs12462273 and rs62107763) and *APOA5* locus (lead SNP rs3741298) affected VK and VE simultaneously (**Figure 3d**).

**Figure 3.**
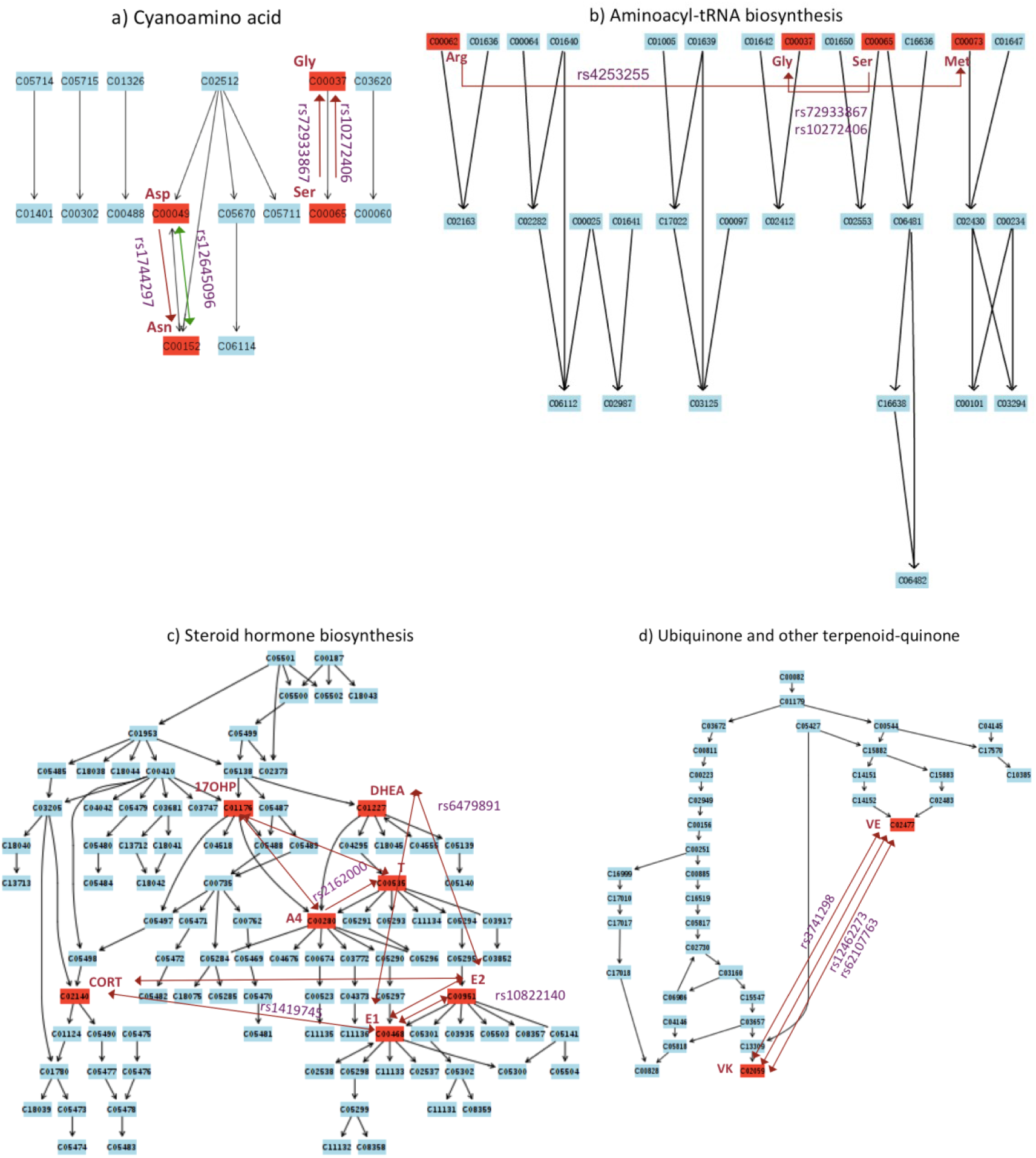
Genetic variants that affect multiple metabolites in the context of KEGG pathway. Shown are SNPs that suggest a significant causal effect in the wald ratio estimates (p<0.05/53). Red lines suggest the same direction of the effect allele while the green line indicates the opposite direction of the effect alleles for the two connected metabolites. Two-edged arrows suggest significant effects of the SNP were observed on both connected metabolites while the single-edged arrows suggest a significant effect on the pointed metabolite. Statistic details of the wald ratio estimates can be found in Table S8. KEGG pathway plot was generated using the online pathway analysis module at https://www.metaboanalyst.ca/.

The KEGG pathways lack information on interactions between trace elements and between metabolites of different categories. Nonetheless, we found that the *CXorf57* (rs72618342), *SERPINA7* (rs1804495), and *MORC4* (rs6616617) loci affected I and Sr levels in the opposite direction (Figure S10). Moreover, the *EGR2-NRBF2* (rs10822140) and *CYP3A5* (rs1419745) loci displayed bi-directional effects between any two hormones in E2, E1, CORT, and Ser (Table S9). Lastly, the *GGH* (rs72658350) locus suggested an effect of VE on Glu level but not the other way around (Table S9, Figure S10).

### Causal maternal metabolites for pregnancy phenotypes and human complex traits

To investigate the newly discovered genetic associations with metabolites, we performed a colocalization analysis for each of the 53 significant loci, using around 100 pregnancy phenotypes from a companion study (Zhu et al., manuscript in submission). Furthermore, we conducted a bi-directional two-sample Mendelian randomization (MR) analysis between the metabolites and 120 phenotypes from the Biobank Japan Project (BBJ), released in the IEU open GWAS project^40^ (see Methods).

The results of the colocalization analysis revealed six loci that demonstrated a strong signal of colocalization between maternal metabolites and pregnancy biomarkers (posterior probabilities of H4, PPH4 > 0.5) (Figure S11). These included shared pleiotropic effects between maternal Mg and serum creatinine (CR) and uric acid (UA) at the AGMAT locus (Figure S11a), vitamin VA and serum creatinine (CR) and pre-albumin (PA) at the GCKR locus (Figure S11b), E1, E2 and the mean platelet volume (MPV) at the EGFR-NRBF2-JMJD1C locus (Figure S11c), 3MHis and urine glucose (GLU_U) at the FBXL20-CDK12 locus (Figure S11d), Fe and mean corpuscular hemoglobin (MCH) at the TST-TMPRSS6 locus (Figure S11e), and I and free thyroxine (FT4) at the SERPINA7 locus (Figure S11f). These findings contribute to new knowledge of pleiotropy between maternal metabolites and biomarker traits during pregnancy.

In the mendelian randomization analysis, after excluding heterogeneity, horizontal pleiotropy, and reverse causality (see Methods), we identified significant causal relationships between seven metabolites and 15 human traits and diseases from BBJ (*P*<0.01) (**Figure 4**). We found that genetically predicted MF5 was associated with increased high-density lipoprotein (HDL) (beta=0.10, 95% CI: 0.04 to 0.16, P= 8.56×10^-4^) and a reduced risk of ischemic stroke (OR=0.84, 95% CI: 0.73 to 0.95, P= 7.07×10^-3^). The discovery of the relation between MF5 and these two traits verified previous MR study and randomized controlled trials (RCT) in the European population^41,42^. Although MF5 protects against ischemic stroke, no causal effect of MF5 on cardiovascular disease such as coronary artery disease was identified in the BBJ (P= 0.62), consistent with the biological function of folate in the nucleic acid synthesis and DNA repair, and medical evidence of folate in nervous system development^43^.

**Figure 4.**
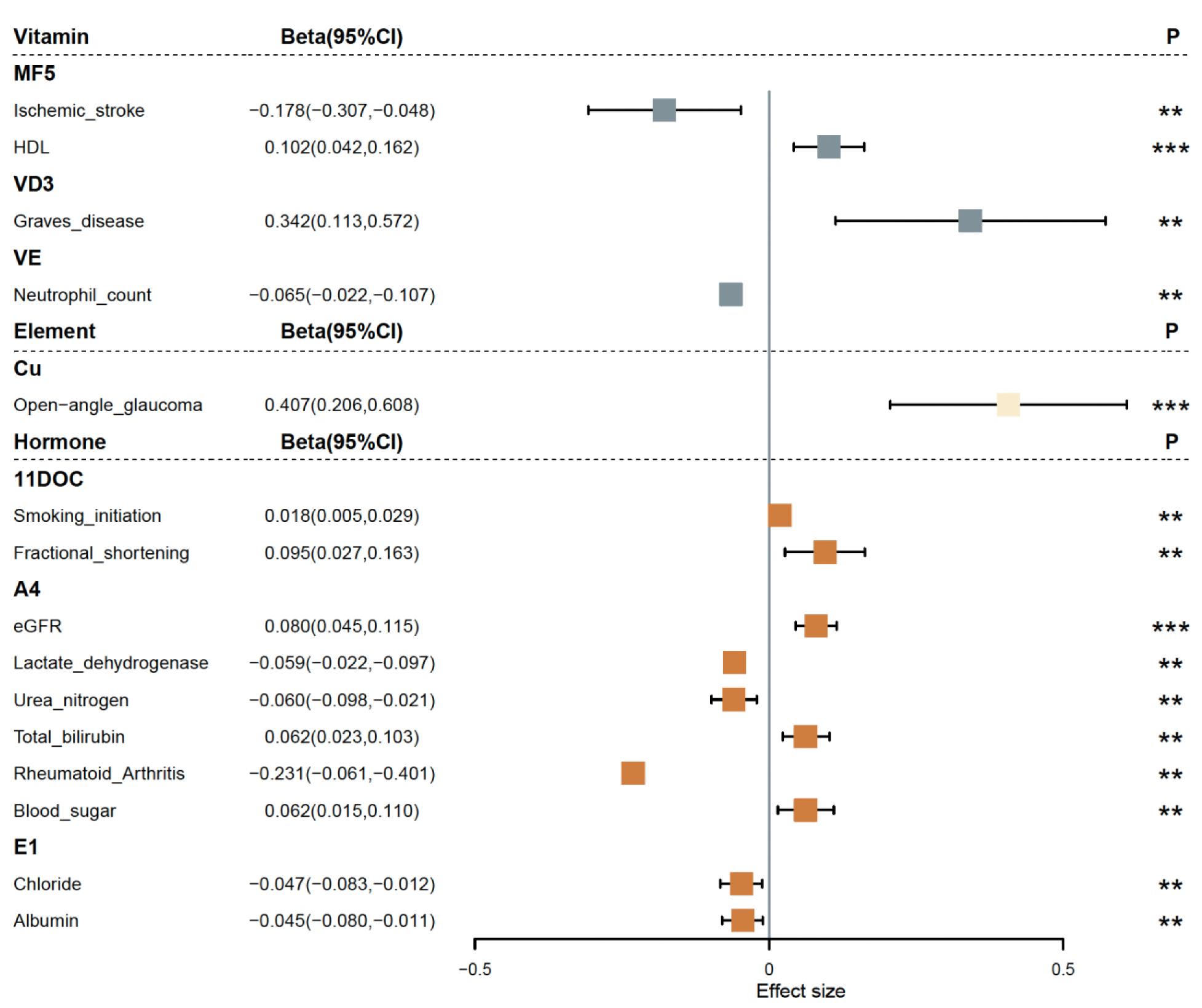
Forest plot displaying the effect of the pregnant metabolites on adult traits by two-sample mendelian randomization. Effects and *P* values were shown if *P*<0.01 based on a Bonferroni correction of five categories of metabolites. The colors of the boxes correspond to the metabolite categories in Figures 1 and 2. The inverse variance weighted method was applied for the MR analysis and only potential causal relationships without horizontal pleiotropic effect (P>0.05) and heterogeneity (P > 0.05) were shown. **: P<0.01; ***:P<0.001

Furthermore, the MR analysis suggested that genetically predicted VD3 was associated with an increased risk of Graves’ disease (GD) (OR=1.40, 95% CI: 1.12 to 1.77, P= 3.41×10^-3^). This differs from observational studies, which suggested that patients with GD were more likely to be deficient in VD compared to the controls, based on a small sample size of less than 100^44^,. However, RCTs did not prove that VD supplementation improves the course of GD^45^. The most recent RCT observed a trend for increased risk of treatment failure and relapse of hyperthyroidism with VD supplementation^46^. An MR analysis in the UK Biobank (UKBB) Europeans suggests no significant associations between VD3 and hyperthyroidism^47^. Therefore, to confirm the effect of VD3 on GD observed in this study, further replication and mechanism validation are required.

Regarding the metal elements, we found that genetically predicted plasma Cu was associated with an increased risk of open-angle glaucoma (OR=1.51, 95% CI: 1.23 to 1.84, P= 7.21×10^-5^).As for the hormones, genetically predicted 11DOC was associated with an increased risk of smoking initiation, consistent with previous observational evidence^48^, and a larger fractional shortening (FS, the size of the ventricle at the end of systole and diastole) derived from the transthoracic echocardiogram (TTE) data. In addition, A4 was associated with a higher level of eGFR, total bilirubin, blood sugar, lower level of lactate dehydrogenase, urea nitrogen, and a decreased risk of rheumatoid arthritis-consistent with reported observational analysis^49^. Lastly, E1 was associated with a lower level of chloride and alumin, which was not reported previously.

## Discussion

We identified a total of 53 genetic associations, including 30 novel genetic associations for 19 metabolites. The consistency of the effect sizes of the associations between the two sequencing technologies demonstrated the robustness of our findings. Notably, our comparison of genetic associations with an independent cohort of non-pregnancy individuals revealed substantial differences in genetic effects on metabolites, especially those of the hormone category. These findings provide valuable insights into human genetics, demonstrating that genetic effects can change or be modified through interactions with internal and external environments, despite DNA stability throughout life. Furthermore, the study also highlights the importance of mechanistic studies to obtain an advanced understanding of the biological mechanisms of pregnancy and reproduction.

We identified pervasive pleiotropic effects in 27 of the 53 metabolite-associated loci (50.94%), which were enriched in nine KEGG pathways, suggesting genetic determination of metabolic flux. Our study emphasizes the importance of multi-trait association analysis rather than analyzing metabolites independently. We also identified novel pleiotropic effects between metabolites of different categories, including the shared effect between I and Sr, between three hormones (E2, E1, CORT) and Ser, and between VE and Glu. Furthermore, analyzing the shared genetic effects between metabolites and a hundred pregnancy phenotypes revealed novel pleiotropy between six metabolites and eight pregnancy biomarkers, unraveling new and complex functions of the metabolites. In our Mendelian randomization analysis of metabolites and 120 phenotypes, we identified potential causal relationships between plasma folate and ischemic stroke, vitamin D3 and Graves’ disease, copper and open-angle glaucoma, and androstenedione and rheumatoid arthritis. These novel findings provide essential information for the development of new therapeutic targets for complex diseases.

Our study has a few limitations. Firstly, due to a lack of birth cohort studies with genetic data in China and East Asians, we were not able to evaluate the potential causal impact of several maternal metabolites on severe pregnancy diseases and adverse birth outcomes reported in observational studies. However, the genetic discoveries obtained from this study made it possible to answer this question in the future, along with the effects in China to build large-scale birth cohorts^51^. Our findings of changing genetic effects during pregnancy, especially in hormones, raise caution for Mendelian randomization studies, which examined the effect of genetically predicted life-long exposures on the outcomes^52^. This will not be a problem for the metabolites that demonstrated consistent genetic effects between the pregnancy and the non-pregnancy status. However, for metabolites that with altering genetic effects such as hormones, how may the pregnancy-specific genetic effect impact late-onset disease will require further investigation. Our report on the potential causal effects of maternal metabolites on human traits and diseases will require more biological validation and replication from independent studies. Lastly, although the current studies represented the largest-scale metabolite genome-wide association studies in East Asians, expanding the sample size and types of metabolites will facilitate a more systematic understanding of biological pathways. Finally, the genetic associations identified from this study are from a single time point in early pregnancy, we are expecting that more metabolite changes will appear in the later stage of gestation. However, the current study provides a first view on the genetic effects on metabolites during the pregnancy period and we have observed substantial changes in genetic effect, suggesting the metabolites already changes in the early period of pregnancy.

This study consistently proves that NIPT sequencing data can be used for medical genetic studies following our previous study introducing the utility of the non-invasive prenatal sequencing data in human genetics^53^. As NIPT sequencing is quickly expanding to more than ten million individuals around the world nowadays, sophisticated study designs and suitable methods will enable the use of this invaluable genetic resource. The methods and knowledge obtained from this study can speed up future efforts from the medical and scientific community to integrate multi-omics data with NIPT data to answer fundamental biological and clinical questions.

## Acknowledgements

We thank all the patients who participated in this study. We would like to particularly thank Professor Anders Albrechtsen from University of Copenhagen and Professor Rasmus Nielsen from University of California, San Francisco for helpful suggestions on the study. We also thank Shujia Huang, Xiao Liu and Fei Wang for their contributions on this study. The study was supported by National Natural Science Foundation of China (31900487, 81830041), Natural Science Foundation of Guangdong Province, China (2017A030306026), Shenzhen Key Laboratory of Genomics (CXB200903110066A) and Guangdong Enterprise Key Laboratory of Human Disease Genomics (2011A060906007), Guangdong Provincial Key Laboratory of Genome Read and Write(2017B030301011) and Guangdong Provincial Academician Workstation of BGI Synthetic Genomics(2017B090904014).

## Author contributions

Conceptualization: Siyang Liu, Xin Jin, Siqi Liu, Liang Lin, Fengxiang Wei, Jianmin Niu, and Rong Wang;

Methodology: Siyang Liu, Yan Li;

Validation: Yan Li, Chao Nie, Fengxia Su, Ya Gao, Fang Chen;

Investigation: Siyang Liu, Jilong Yao, Liang Lin, Xianmei Lan, Linlin Wu, Nannan Kong, Yuqing Deng, Yan Li, Jiansheng Xie, Zilong Li, Likuan Xiong, Jinghui Ren, Xuemei Qiu, Weihua Zhao, Ya Gao, Yuanqing Chen, Fengxia Su, Yun Zhou, Weiqiao Rao, Jing Zhang, Guixue Hou, Liping Huang, Xinhong Liu, Chao Nie, Liqiong Luo, Zengyou Liu, Fang Chen, Shengmou Lin, Lijian Zhao, Dan Jiang, Ye Yin, Xun Xu, Jian Wang, Huanming Yang, Rong Wang, Fengxiang Wei, Xin Jin, Siqi Liu;

Formal Analysis: Siyang Liu, Xianmei Lan, Zilong Li, Nannan Kong, Yan Li; Data Curation, Nannan Kong, Yuan Wang, Xiaoxia Wu;

Writing-Original Draft: Siyang Liu;

Writing-Review & Editing: Siyang Liu, Siqi Liu;

Visualization: Siyang Liu, Xianmei Lan, Zilong Li, Nannan Kong; Supervision: Siqi Liu, Jian Wang, Huangming Yang;

Project Administration: Liang Lin; Funding Acquisition: Xin Jin, Siyang Liu;

## Methods

### Participant recruitment and plasma sample preparation

All the 34, 394 participants were recruited via the non-invasive fetal trisomy test and the pregnancy nutrition program between year 2017 and 2018. They underwent pretest counseling and filled in informed written consent before blood sampling. 16The study was reviewed and approved by the Institutional Review Board of BGI (BGI-IRB21184) in strict compliance with regulations regarding ethical considerations and personal data protection.

5ug peripheral whole blood was drawn from each of the participant and stored in the EDTA anticoagulant tubes to avoid hemolysis. The plasma was obtained by centrifugation (3000 rpm, 10 min) and was preserved at -80°C until assay. As for amino acid extraction, 40 μL plasma was mixed with 20 μL stable-isotope labeled internal standard (IS) in sulfosalicylic acid to precipitate proteins, followed by vortex and centrifugation (4000 rpm, 4°C, 20 min). Regarding hormones, 250 μL plasma mixed with 205 μL IS solutions were filtered through solid-phase extraction (SPE), followed by washing with 25% methanol and eluting with 100% dichloromethane.

The elution was evaporated by nitrogen and was reconstituted in 25% methanol. For vitamin extraction, approximately 200 μL plasma were mixed with 600 μL methanol/acetic acid/IS for water-soluble vitamins (WSV), or with 1000 μL methanol/acetonitrile/IS for fat-soluble vitamins (FSV), followed by vortex and centrifugation (4000 g, 4°C, 20 min). The supernatants were evaporated by nitrogen and were reconstituted by either 60 μL deionized water (WSV) or 80 μL 80% acetonitrile (FSV). As regards metal elements, 100 μL plasma were mixed with 400 μL nitric acid/hydrogen peroxide for decomposition at 105°C for 3 hours. The decomposed products were cooled down and delivered to mass spectrometry after dilution with deionized water.

### Quantitative measurement of metabolites using mass spectrometry

The metabolites in blood were targeted and quantified using liquid chromatography coupled with triple quadrupole mass spectrometry (LC MS/MS), including ACQUITY UPLC I-Class (Waters) mounted with C18 column, Triple Quad 5500 (Sciex) and Xevo TQ-S (Waters). The MS/MS spectra corresponding to metabolites were acquired at positive ion mode with multiple reaction monitoring scans. The metal elements were measured by inductively coupled plasma mass spectrometry (ICP MS/MS), i.e. 7700x ICP-MS (Agilent). The mass spectra acquired were processed with MultiQuant (V. 3.0.2, Sciex) for amino acids, hormones and FSV, MassLynx (V. 4.1, Waters) for WSV and MassHunter (V. B.01.03, Agilent) for metal elements. The calibration curves were implemented with stable isotope-labeled compounds as internal standards. The accuracy of quality controls (QCs) with isotope-labeled IS was managed approximately every 15 samples to ensure the inter-batch stability.

### Sequencing assays

Details of the sequencing protocol were published previously in ^54^. In brief, within 8h of blood collection, plasma was extracted from whole blood after two turns of centrifugation. The plasma samples were subsequently subjected to library construction, sample quality control and 36-cycle single-end multiplex sequencing on BlackBird or BGI-seq500 platform. The reads were trimmed to 35bp before bioinformatic analysis. Filtering of poor quality reads was carried out using SOAPnuke (https://github.com/BGI-flexlab/SOAPnuke). A read was removed if it contained more than 30% low quality bases (Q<=2) or N bases. In general, each participant was whole-genome sequenced to 5-10 million cleaned reads, representing a sequencing depth around 0.06x -0.1x.

### Correlation between metabolite concentrations

We have investigated the correlation between metabolite concentrations in two aspects. First, we estimated the spearman correlation coefficient between the phenotypes. Second, we applied the correlation model implemented in LD score correlation (LDSC) to compute the genetic correlation between any two of the metabolites (Bulik-Sullivan et al., 2015). Correlation was visualized as a heatmap using R.

### Genotype imputation

We employed STITCH (version 1.2.7) ^55^ to impute genotype probabilities for the BB and Seq500 individuals in a five-megabase window with a 250K buffer assuming 40 ancestral haplotypes, respectively. Allele frequency information from the Chinese population (CHB+CHS+CDX, N=301) in the 1KG impute2 reference panel was used for the initial values for the EM optimization of the model parameters. The imputed variants included 8.16 million known polymorphic sites in 22 autosomal chromosomes and chrX with a 1KG East Asian allele frequency >=0.01. All the loci recorded in the GWAS catalog are also included for imputation. For each of the imputed sites, there is an IMPUTE2-style info score (Marchini et al., 2007) and a P-value for violation of Hardy Weinberg equilibrium (HWE-pvalue in short) (Wigginton et al., 2005). We used info score greater than 0.4, and minor allele frequency greater than 0.01 as filtration threshold to obtain the significantly associated variants. Imputation accuracies were estimated using Pearson’s R2 between the true genotype from high-coverage WGS of 50 participants (40x) and the imputed genotype dosage.

### Genome-wide association analysis with PLINK

Since the variance regression approach assumes that the trait values follow multivariate normal distribution, we performed a rank transformation of the metabolite raw values using the empirical normal quantile transformation approach after removal of outliners in the raw value distribution^56^. We then applied the linear regression model implemented in Plink v1.9 to detect the associations between the imputed genotype dosage and the transformed values of the metabolites ^57^. The covariates in the regression model include maternal age, gestational week upon sampling, the top three principal components of PCA and the inferred gender of the fetus computed from the NIPT data. The regression analysis was performed for black bird and Seq500 sequencing data independently that produced effect size, standard errors, number of effect individuals and p-value for each of the imputed sites.

### Meta-analysis and multi-trait genome-wide association test using the MTAG approach

We applied the inverse variance based approach implemented in METAL to perform the meta-analysis integrating the effects from BB and Seq500 regression outcomes ^26^. Since many metabolites may have shared genetic correlation, we applied the multi-trait genome-wide association studies based on the metal summary statistics using the MTAG approach ^27^. The resulting beta, p-value and standard errors were used for downstream analysis. During the MTAG analysis, we noticed that the time and memory that required for performing multi-trait genome wide association studies using all the 84 metabolites is too consuming due to very large Omega and Sigma matrices and we cannot finish the computation successfully in the end. After a personal communication with the MTAG author, we performed the MTAG analysis for each amino acid (N=37), element (N=24), hormone (N=13) and vitamin (N=10), respectively.

### Identification of independent significant loci

After learning the effect size and the p-value from the meta-analysis, we defined independent loci as significant variants clustered in a 1Mbp window. The lead SNP was defined as the SNP in the 1Mbp window that has the smallest p-value. We didn’t observe multiple independent signals in one locus after performing a conditional test using Plink v1.9. Furthermore, locuszoom was applied to visualize the loci ^58^. The reported loci were determined from the conditional test after the single marker analysis using a significance threshold P value ≤ 5 x 10^-8^. Since the genome-wide association study was performed on the quantile-transformed metabolite value (β_nor_), we applied the following formula β = β_nor_ * sd_pheno_ to obtain the effect on real metabolite level where sd_pheno_ refers to the standard deviation of values of certain phenotype. Both β_nor_ and β were reported in Table 1 and Table S3.

### SNP heritability

The genomic inflation factor λ_GC_, the heritability, the intercept, and ratio using the LD score regression approach based on the summary statistics^35^. 12 out of the 84 metabolites display negative heritability. After personal communication with the LDSC author, this may be due to a lack of power for those traits because of small sample size. We set those metabolites with negative heritability as zero in Figure S8 and Table S7. The UK Biobank heritability was obtained from https://nealelab.github.io/UKBB_ldsc/.

### Comparison with the non-pregnancy population

1,553 participants (Female N=642, Male N=911) from Shenzhen local area were recruited. Written form of consent was signed by each individual. The white blood cell of each individual were whole-genome sequenced to around 30x using the BGI-seq500 platform. A total of the 81 out of the 84 metabolites were investigated for each individual using the same protocol for the pregnancy study that the amino acids, vitamins and hormones were assayed using plasma while the elements were assayed using the whole blood. The three metabolites not investigated were I, Ba and DOC. WGS data were aligned and variants called by the Picard (http://picard.sourceforge.net), BWA ^59^ and GATK v3.8 best practice ^60^pipeline. SNPs with mapping quality greater than 40, sequencing depth greater than 4, variant quality greater than 2.0, Phred score of Fisher’s test p-value for stand bias smaller than 60.0, Haplotype score smaller than 13.0 and distance of alternative allele from the end of reads greater than 8.0 were kept for following analyses. We removed SNPs deviating from Hardy-Weinberg (P-value < 1×10-5), markers with more than 1% missing genotype data and variants with smaller than 1% minor allele frequencies. Individuals with heterozygosity greater than there standard deviations were excluded. One individual among relatives within 3^rd^ degree of relationship was randomly selected to keep in the clean data set. PCA was performed to investigate population stratification. No clean sub-cluster was observed. Typical north to south Grandaunt was demonstrated by the first principal component. Linear regression adding individual sex and top two principal components as covariates was performed for each of the significant locus identified from the pregnancy study. We also performed regression using only the female individuals adjusting top two PCs. No inflation was observed in this analysis (λ_GC_ ∼ 1).

### KEGG and pleiotropy analysis

To investigate the pleiotropy among metabolites, for each of the 53 significant loci (P value ≤ 5 x 10^-8^), we visualized the effect and *P* value for the lead SNP in a forest plot (**Figure S10**). Strong pleiotropy was defined as the observation of a lead SNP that affects at least two metabolites with a significance level of 10^-3^. To further understand the relationship between the SNP variants and multiple metabolites, we conducted a wald ratio estimation implemented in the tsls package in R for each of the 27 variants that suggest pleiotropy. We performed a total of 54 tests, corresponding to a p-value less than 0.001 after the Bonferroni correction. We summarized and visualized the pleiotropic correlation in the KEGG pathway using the metaboanalyst website (https://www.metaboanalyst.ca/).

### Colocalization analysis

To investigate whether the same genetic variants may drive the associations with metabolites and the pregnancy phenotypes reported in a companion paper, we undertook a colocalization analysis. We applied a stringent Bayesian analysis implemented in the coloc R package^61^z with default parameters to estimate the posterior probability (PP) that the metabolites and the pregnancy phenotypes shared a single causal SNP at the locus. The SNPs in the 1[Mb range of the tested instrumental variable that have MAF over 0.05 were used for analysis.Metabolites with a PPH4 > 0.5 (posterior probabilities of two traits sharing one causal SNP) were considered to be colocalized.

### Two-sample Mendelian randomization between metabolites and 120 traits from the Biobank of Japan

To investigate the potential causal impact of the metabolite on human traits or diseases, we performed Mendelian randomization analyses using four methods: the inverse variance weighted (IVW) method, the MR Egger method, the weighted median method, and the weighted mode method implemented in the TwoSampleMR R package^62^. The metabolites that passed the correction threshold *P*<0.01 were retained for pleiotropy and heterogeneity evaluation. Cochran’s Q statistic was used to examine the heterogeneity of the IVs. Horizontal pleiotropy was evaluated using the MR Egger approach with a return of intercept values. MR Steiger test^63^ was applied to test the directionality of the effect. Only potential causal effects passing the pleiotropy, directionality, and heterogeneity tests were reported in our study (*P*>0.05).

## Data availability

The summary statistics of the metal meta-analysis outcome and the MTAG outcome have been deposited into CNGB Sequence Archive 22 of CNGBdb23 with accession number CNP0003025.

