## SupplementaryMaterial for "Multi-trait genome-wide association study in 34,394 Chinese women reveals the genetic architecture of plasma metabolites during pregnancy"

### Supplementary Figures

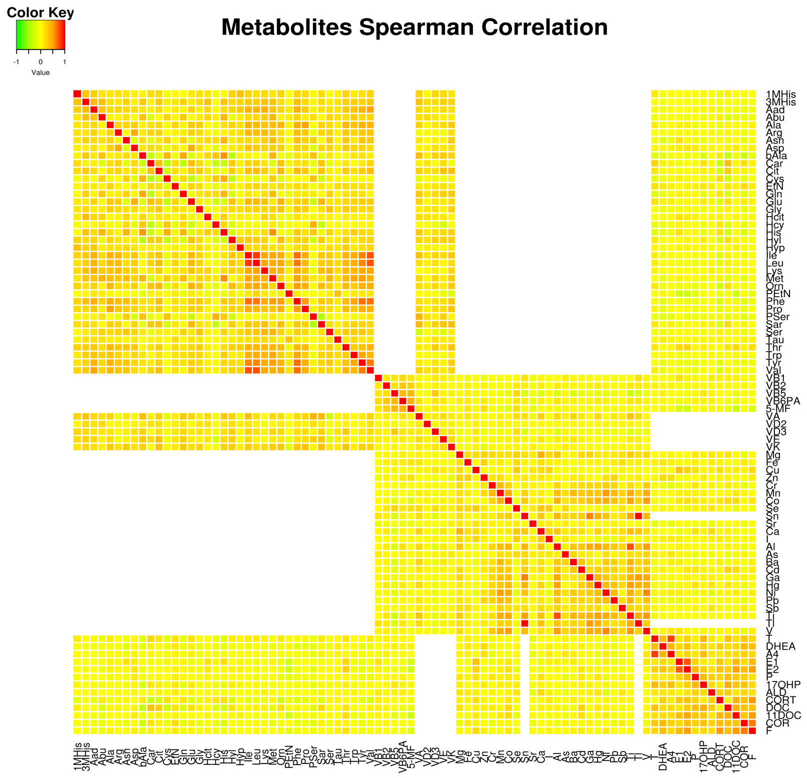

**Figure S1. Spearman pairwise correlation for metabolites**
The spearman pairwise correlation for all 84 metabolites is demonstrated. Negative values are in green, positive values are in red and missing values are colored in the blank.

### *Amino Acids*

#### 3MHis

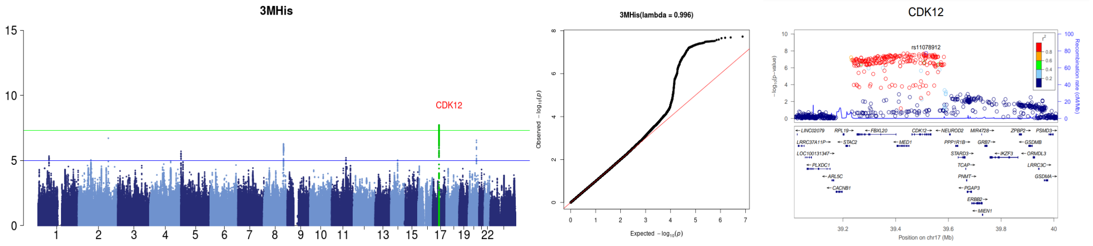

#### Aad

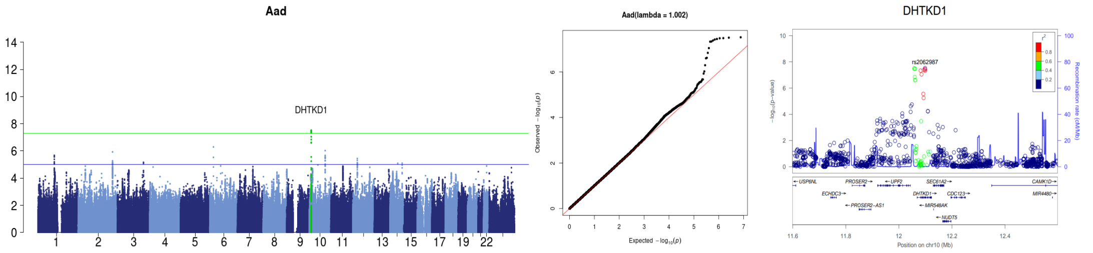

#### Arg

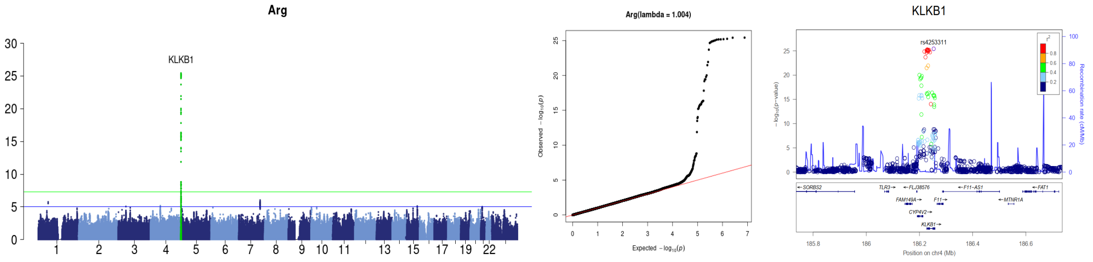

#### Asn

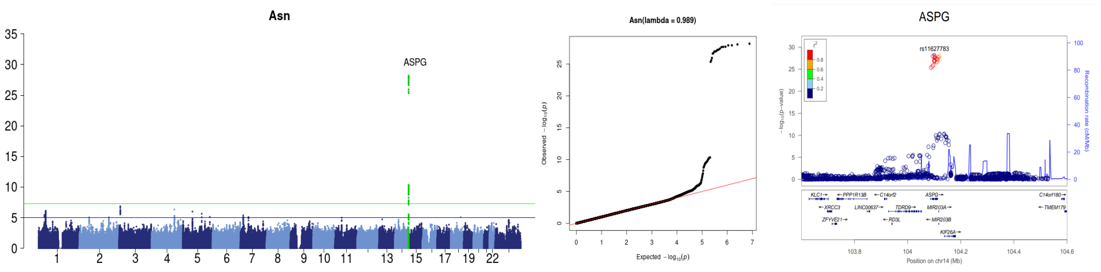

#### Asp

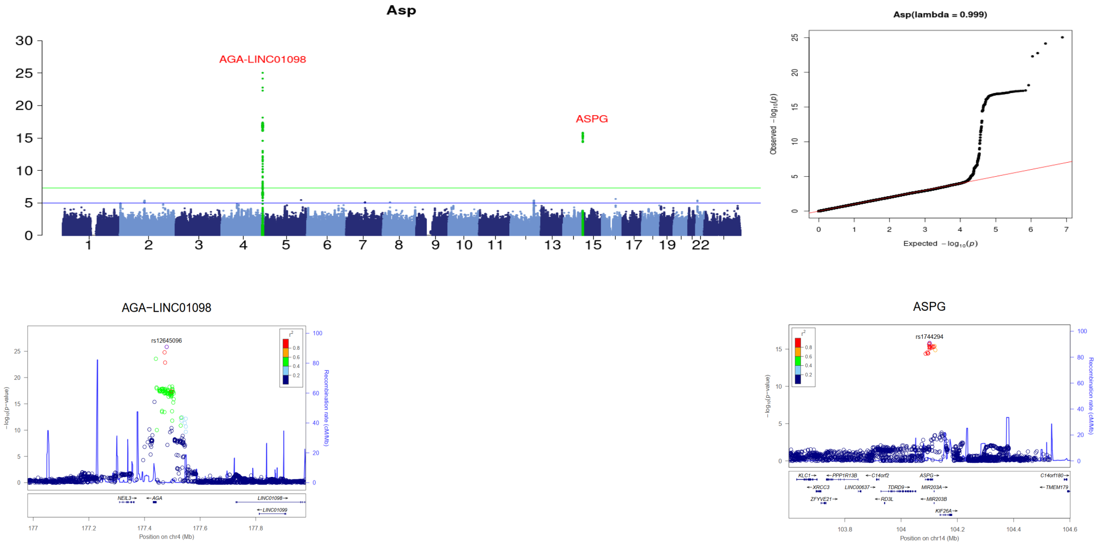
Asp
AGA-LINC01098
ASPG
Asp(lambda = 0.999)
AGA−LINC01098
rs12645096
Position on chr4 (Mb)
ASPG
rs1744294
Position on chr14 (Mb)

### bAla

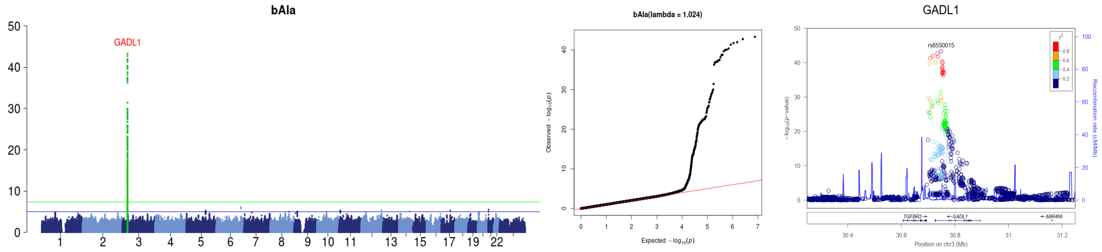
bAla
GADL1
bAla(lambda = 1.024)
GADL1
rs6550015
Position on chr3 (Mb)

### Cit

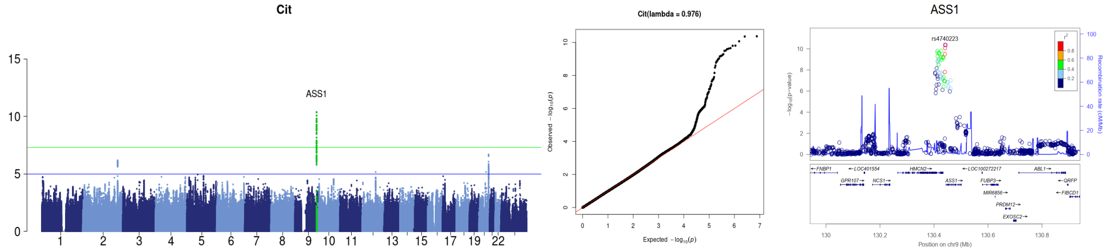
Cit
ASS1
Cit(lambda = 0.976)
ASS1
rs4740223
Position on chr9 (Mb)

### Gly

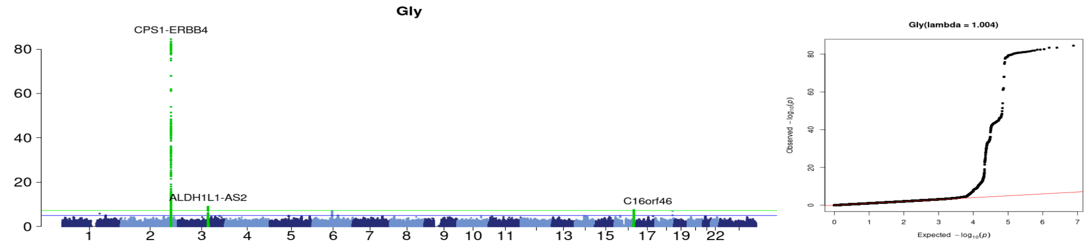
Gly
CPS1-ERBB4
ALDH1L1-AS2
C16orf46
Gly(lambda = 1.004)

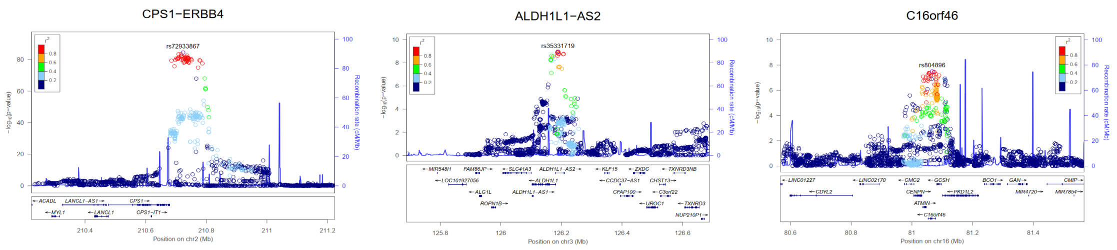

### Hcy

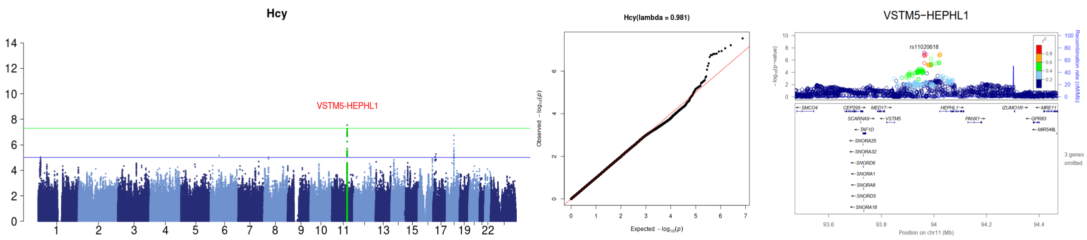

### Hyl

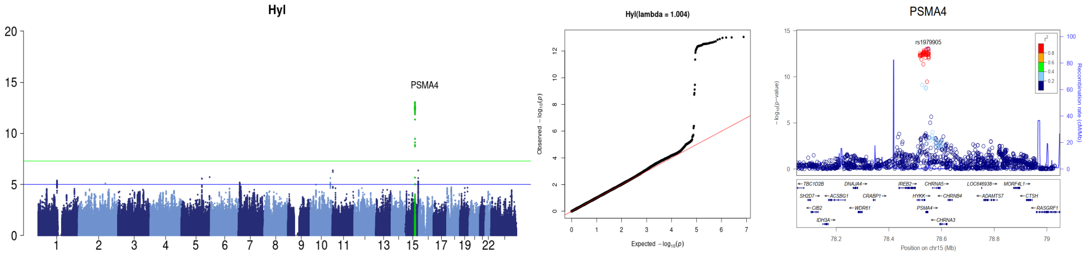

### Sar

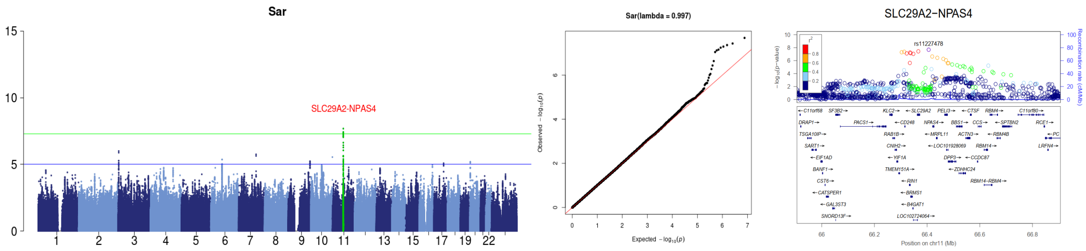

### Ser

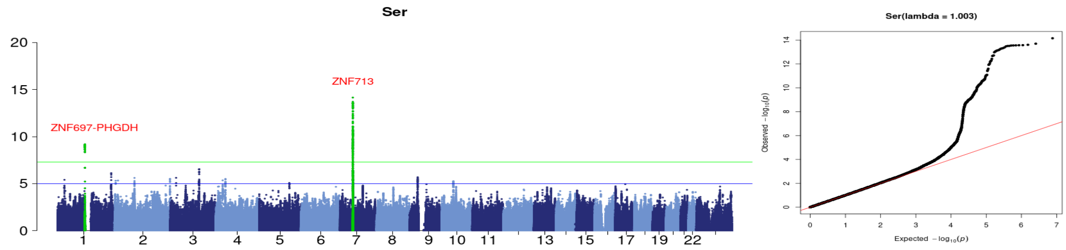

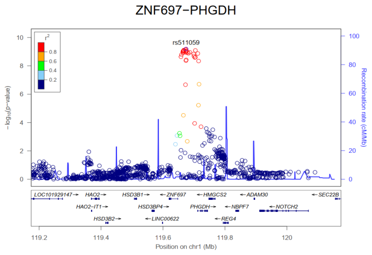

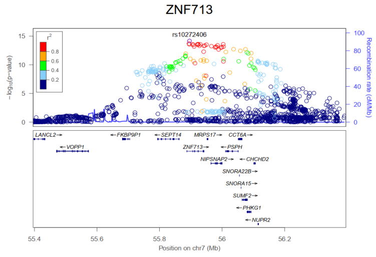

### Tyr

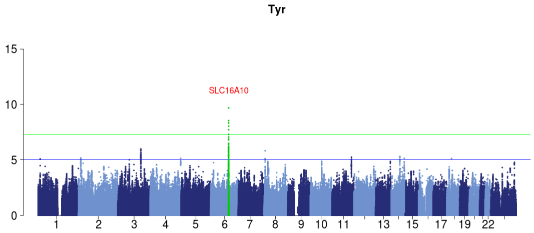

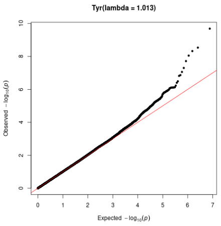

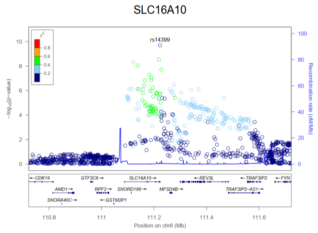

### Val

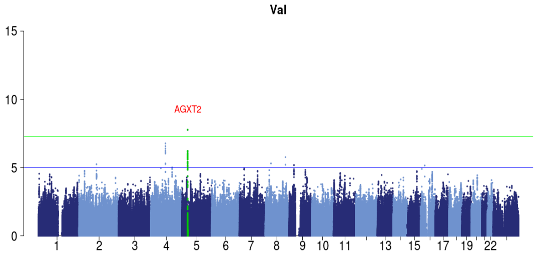

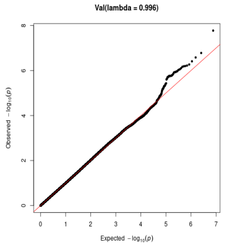

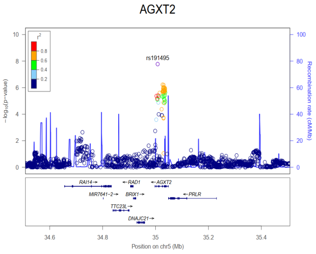

**Figure S2. Manhattan, quantile-quantile, and locuszoom plots for all the 14 amino acids that consist of at least one significant loci**

#### *Vitamins*

## MF5

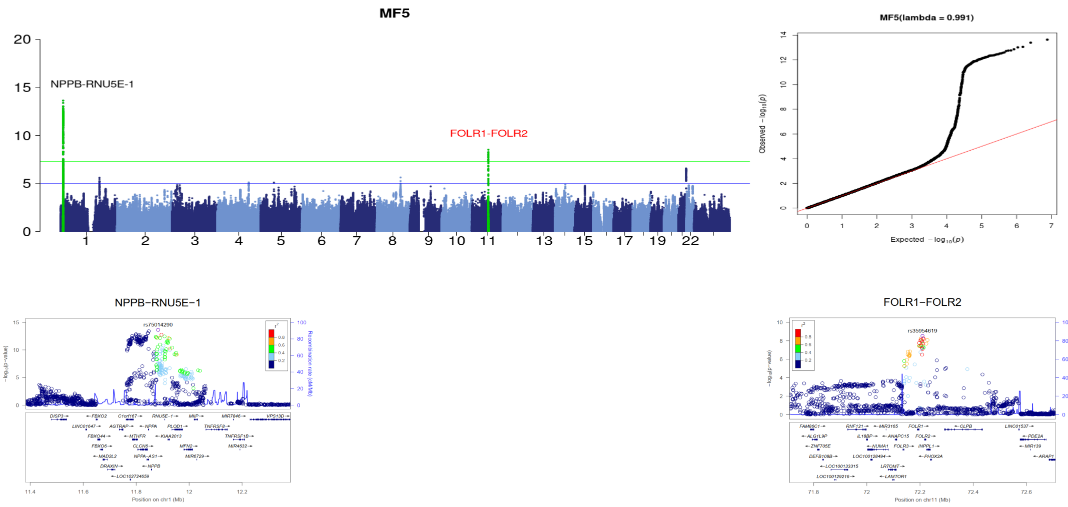

## VA

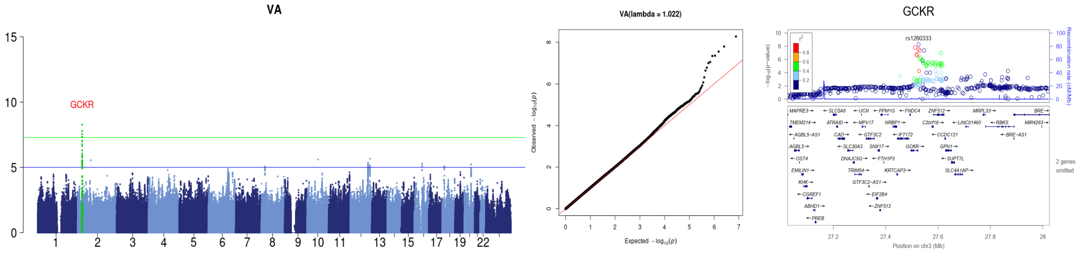

## VB2

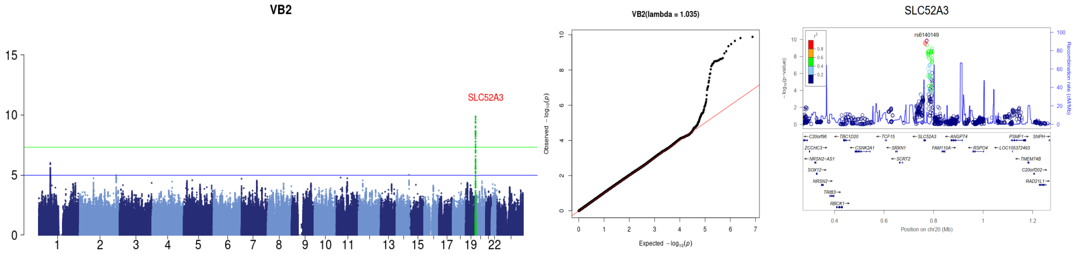

## VB5

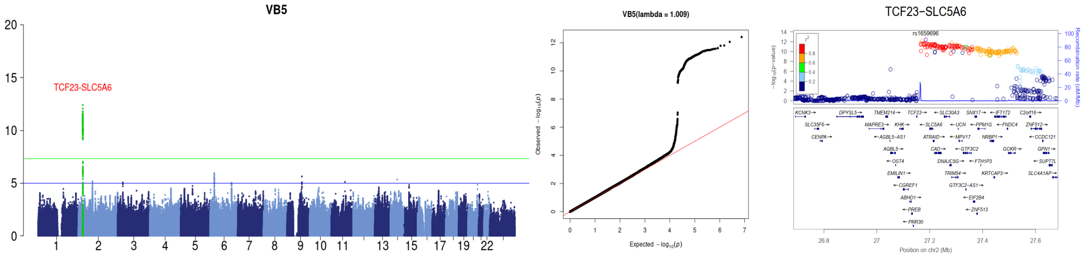

## VD3

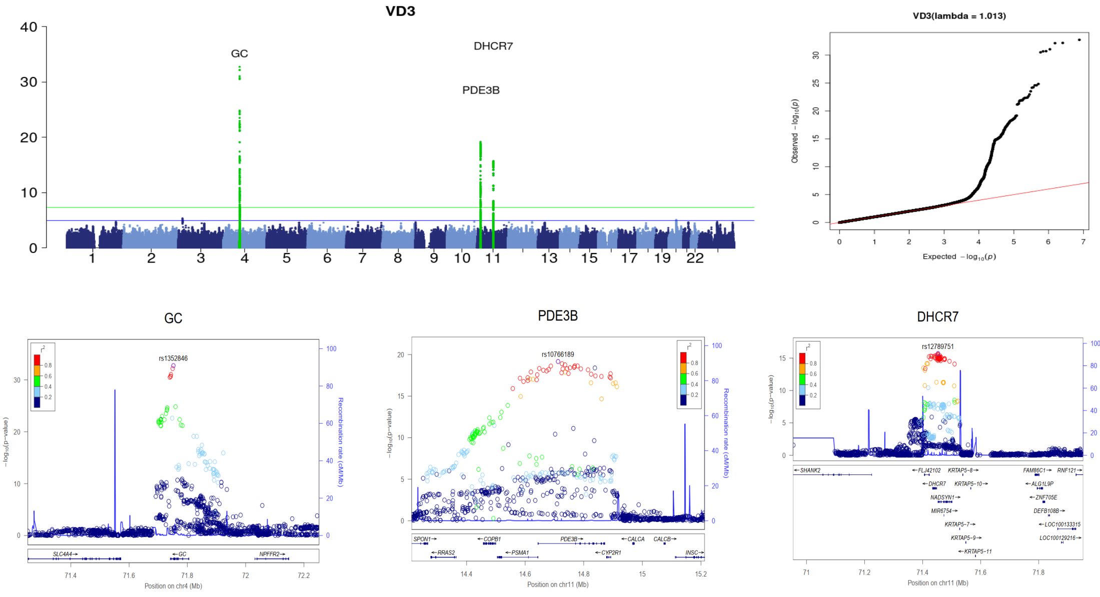

# VE

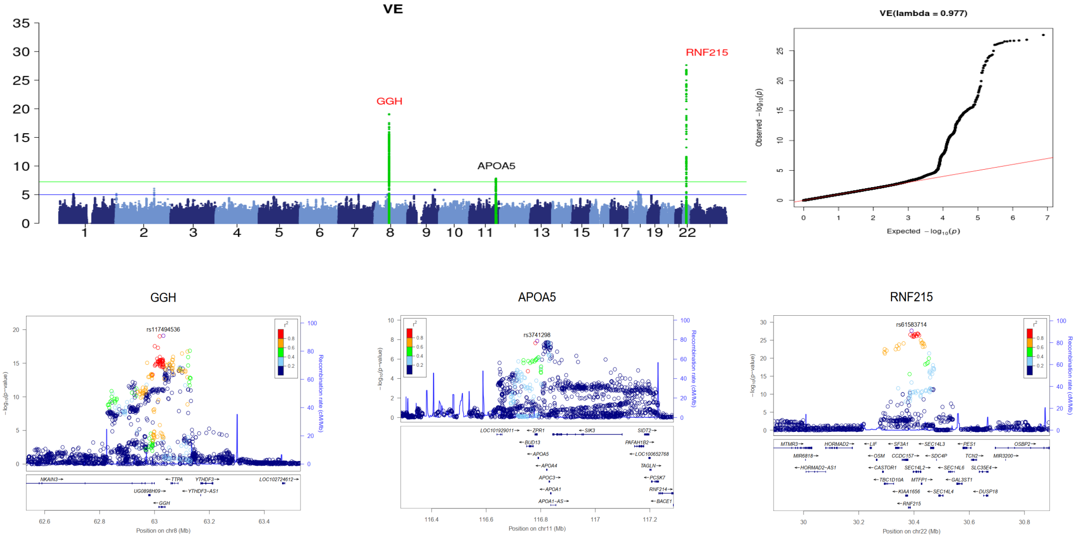

# VK

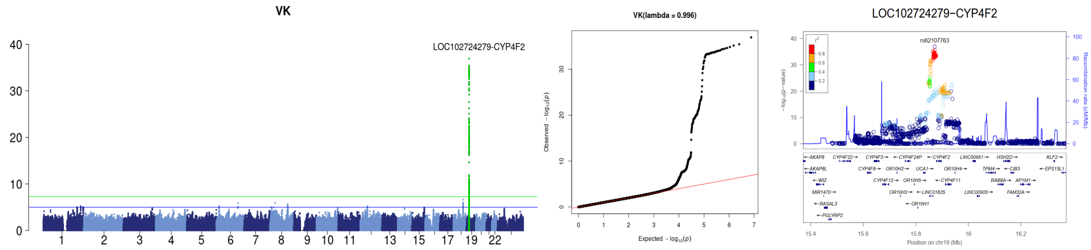

**Figure S3. Manhattan, quantile-quantile, and locuszoom plots for 7 hormones that consist of at least one significant locus**

#### *Elements*

## Ba

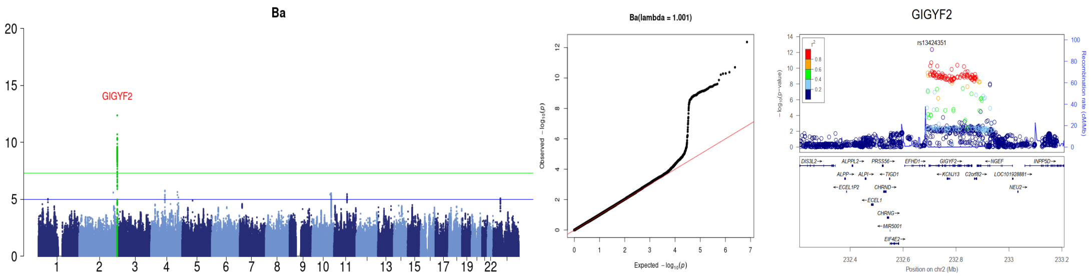

## Cu

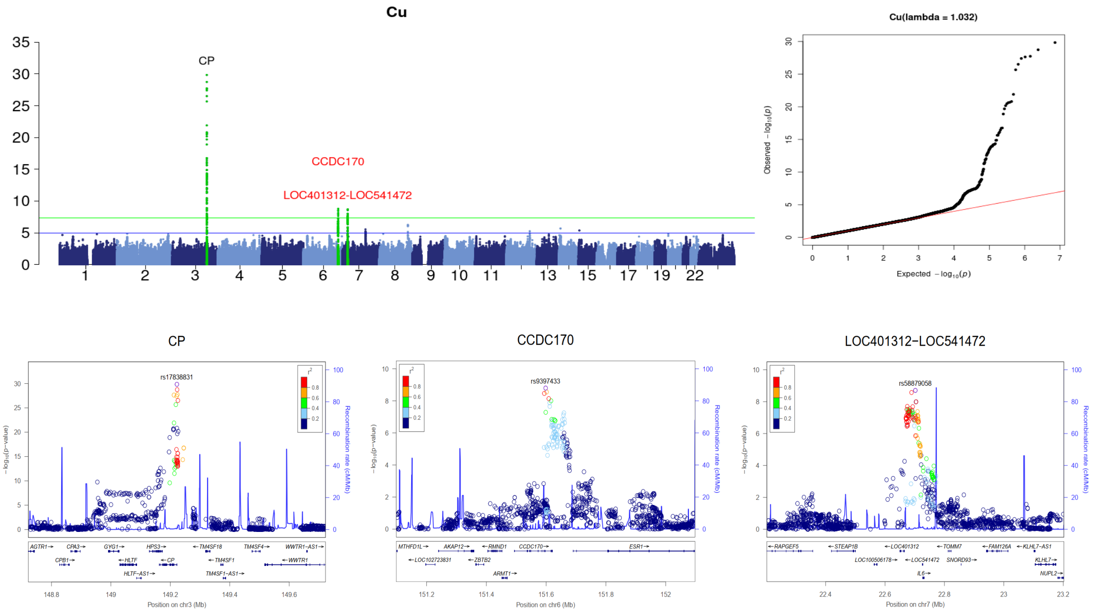

## Fe

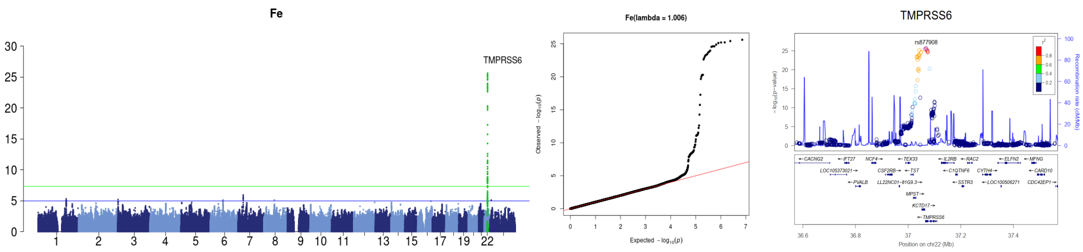

## I

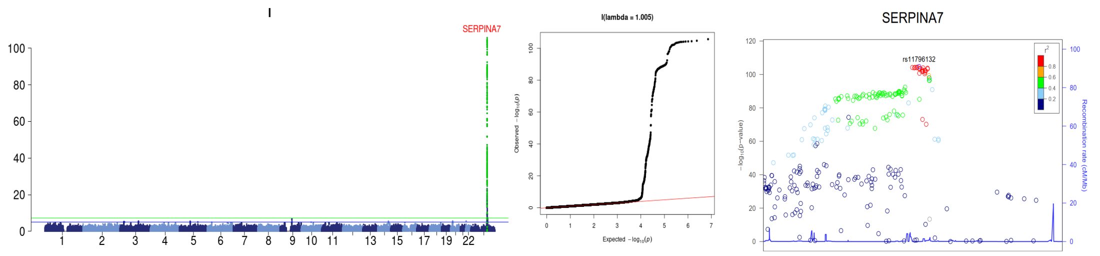
I
SERPINA7
I(lambda = 1.005)
SERPINA7
rs11796132

# Mg

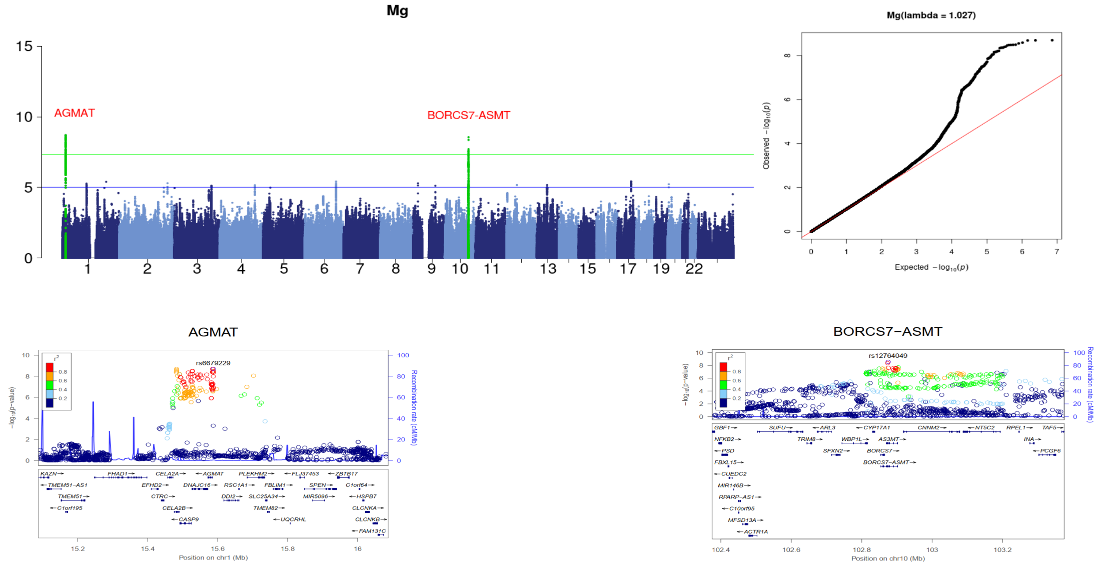
Mg
AGMAT
BORCS7-ASMT
Mg(lambda = 1.027)
AGMAT
rs6679229
BORCS7−ASMT
rs12764049

# Se

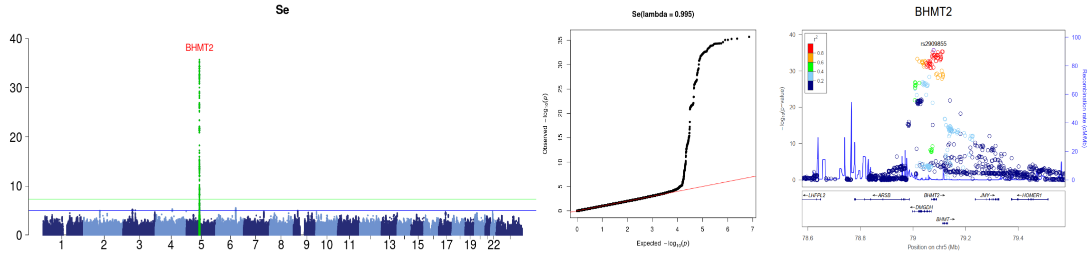
Se
BHMT2
Se(lambda = 0.995)
BHMT2
rs2909855

# Sr

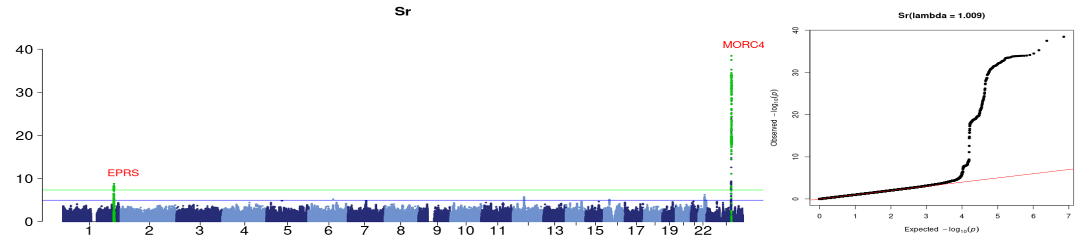
Sr
MORC4
EPRS
Sr(lambda = 1.009)

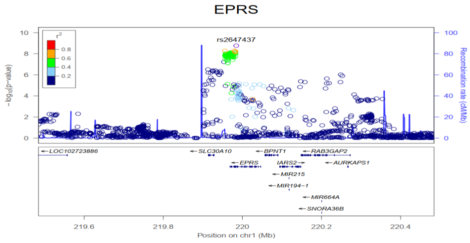

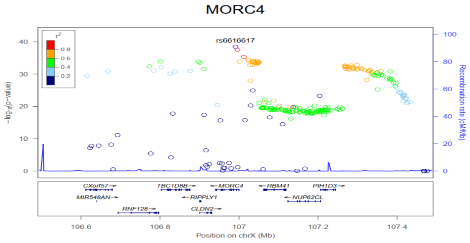

**Figure S4. Manhattan, quantile-quantile, and locuszoom plots for 7 metal elements that consist of at least one significant loci**

### *Hormone*

## A4

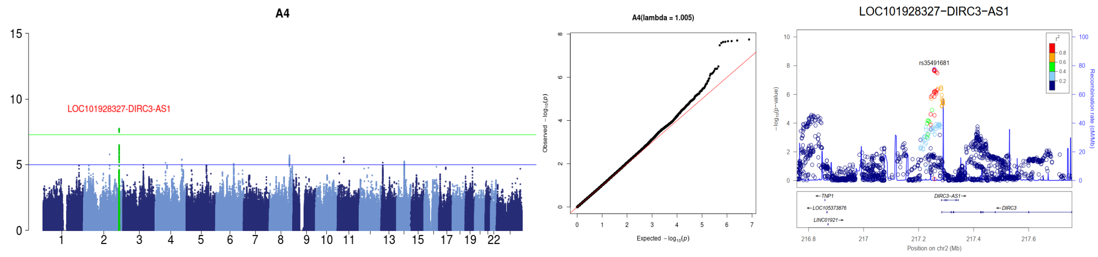

#### DOC

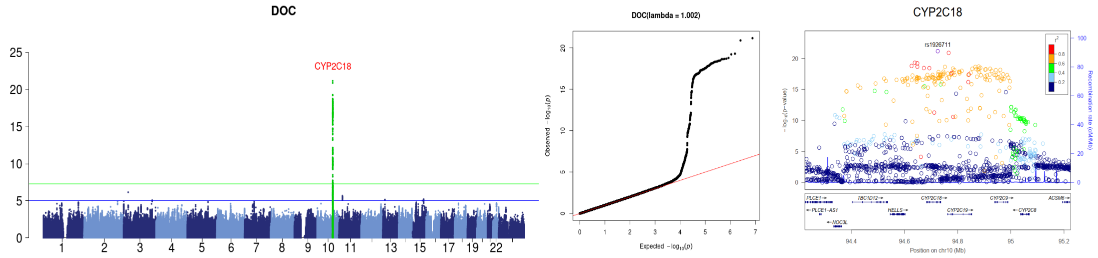

## E1

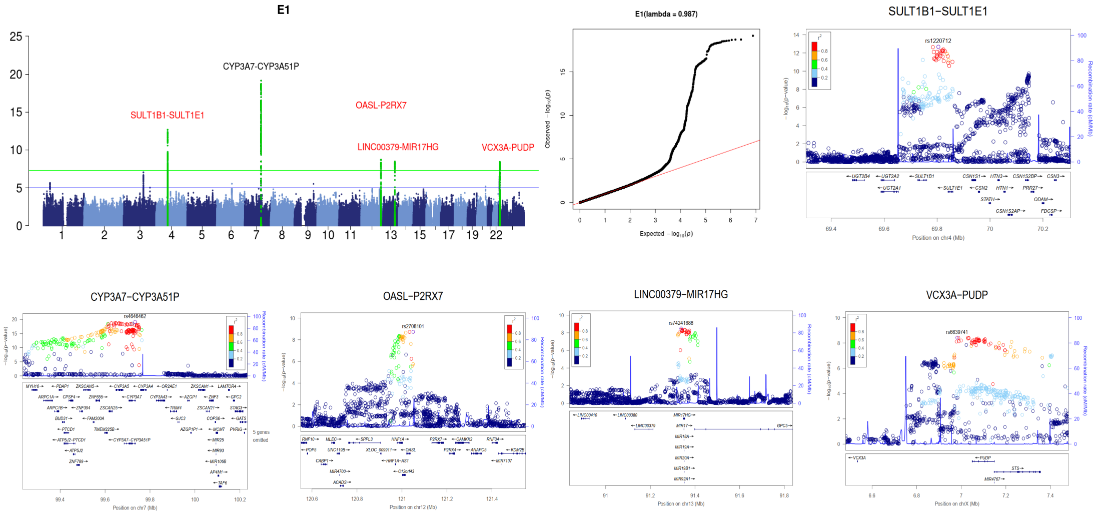

## E2

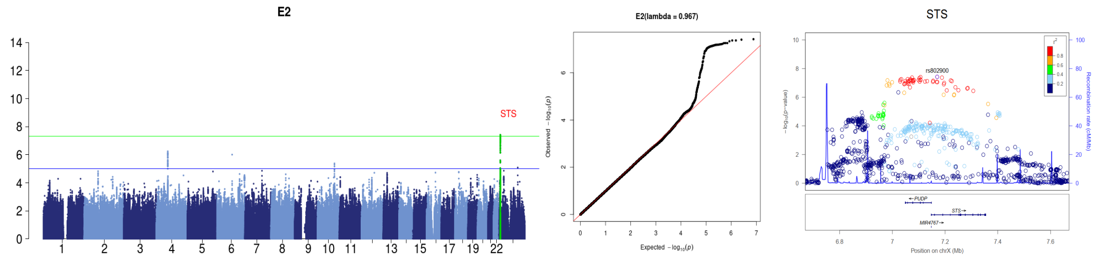

### 11DOC

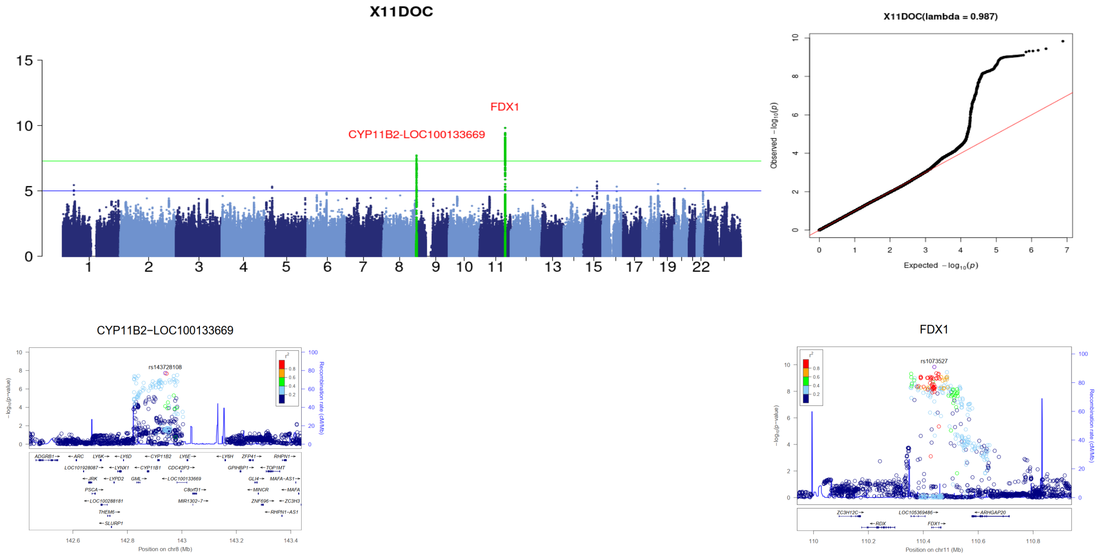

### 17OHP

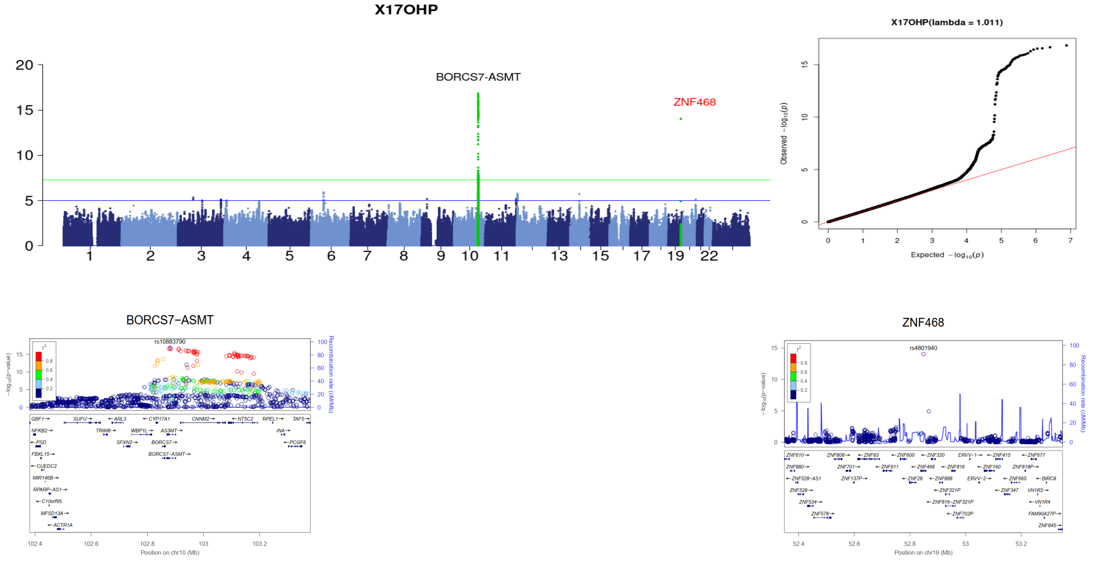

**Figure S5. Manhattan, quantile-quantile, and locuszoom plots for 5 hormones that consist of at least one significant locus**

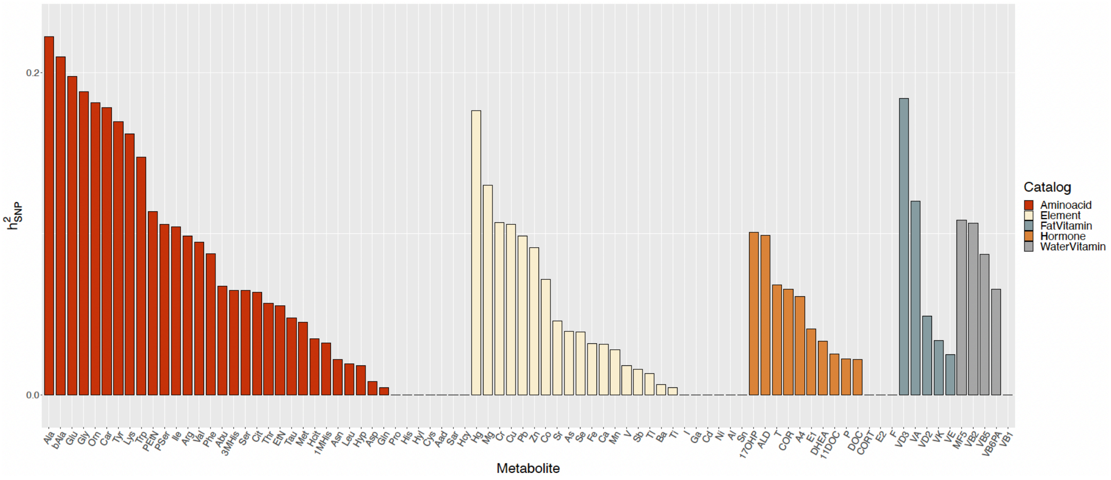

**Figure S6. SNP heritability of the metabolite level in maternal plasma**

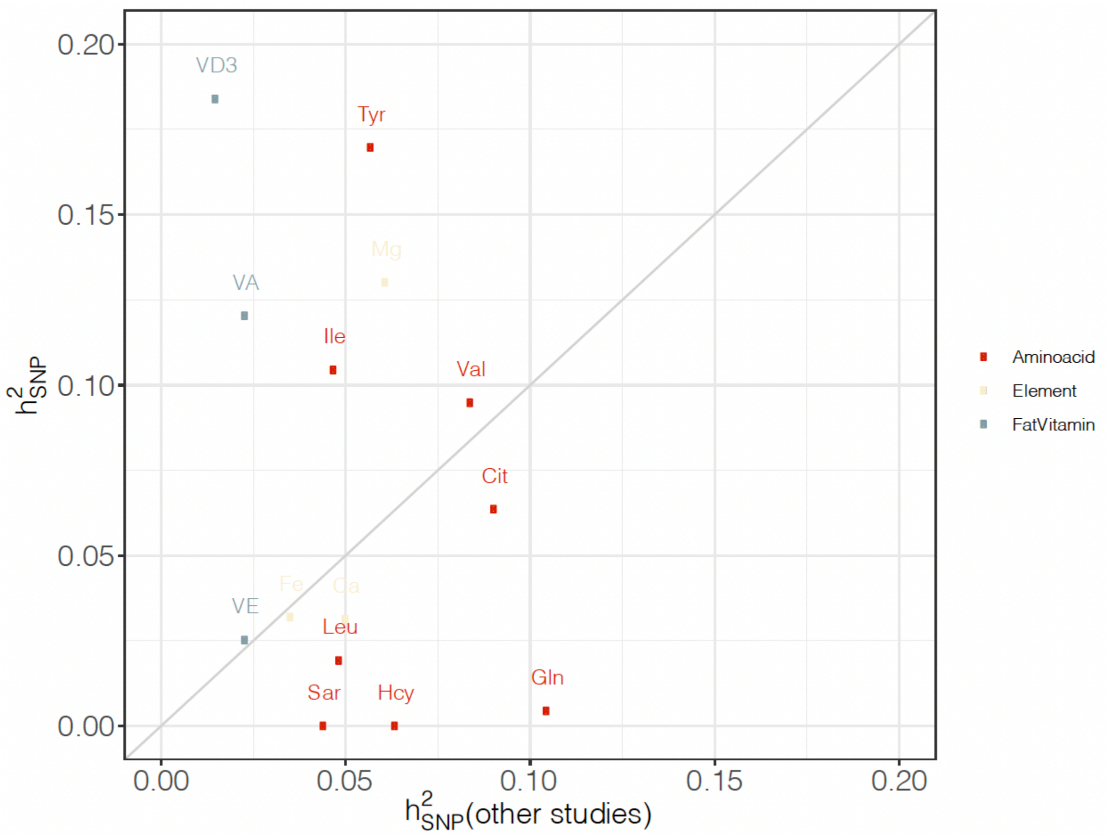

**Figure S7. Comparison of the heritability between the previous study and our study**

Known heritability refers to the heritability estimation from reports of Kettunen et al., 2016 and Neale's lab (Table S6). Only metabolites with previously known heritability were shown in this plot.

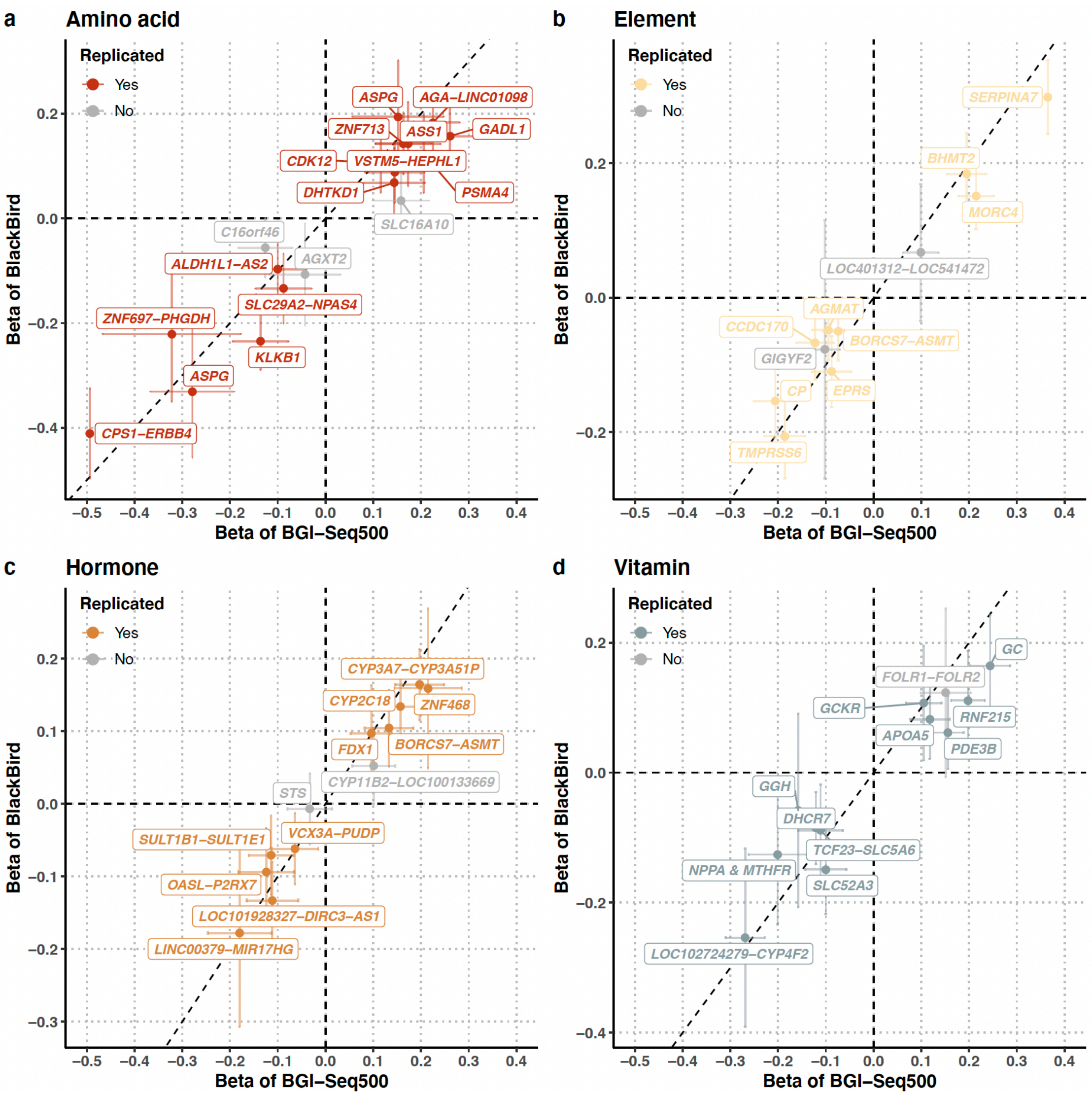

**Figure S8. Comparison of effect size between two sequencing technologies**
Comparison of the effect size of the 53 significant loci between the BGI-Seq500 sequencing and the BlackBird sequencing platform. A locus was denoted as replicated when it has the same direction of effect and p-value smaller than 0.05 in both gwas analysis from both sequencing technologies.

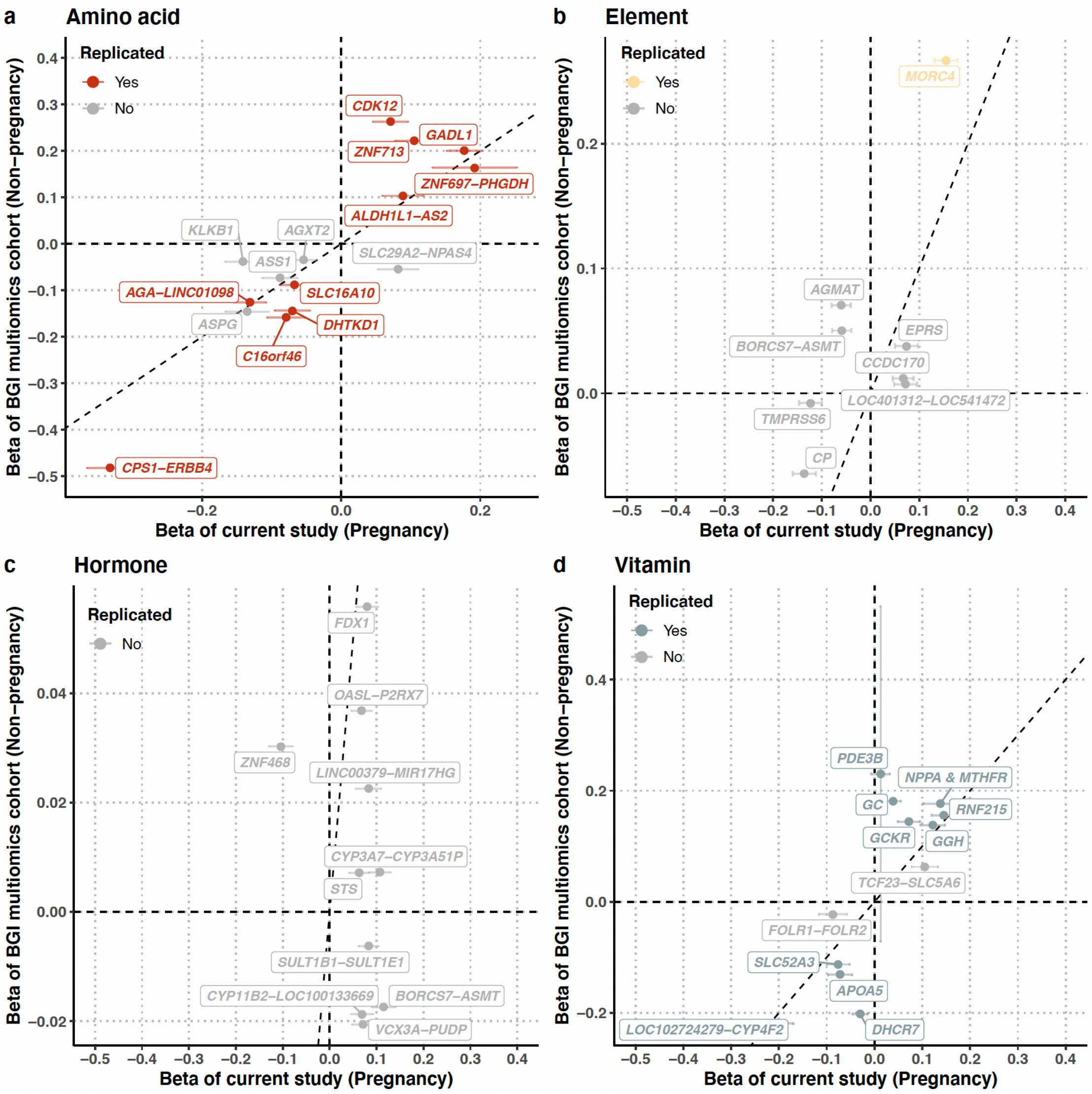

**Figure S9. Comparison of effect size with a non-pregnancy cohort**

Comparison of the effect size of the 53 significant loci between the current study and a non-pregnancy cohort (BGI multiomics cohort). A locus was denoted as replicated when it has the same direction of effect and p-value smaller than 0.05 in both gwas analysis from both sequencing technologies.

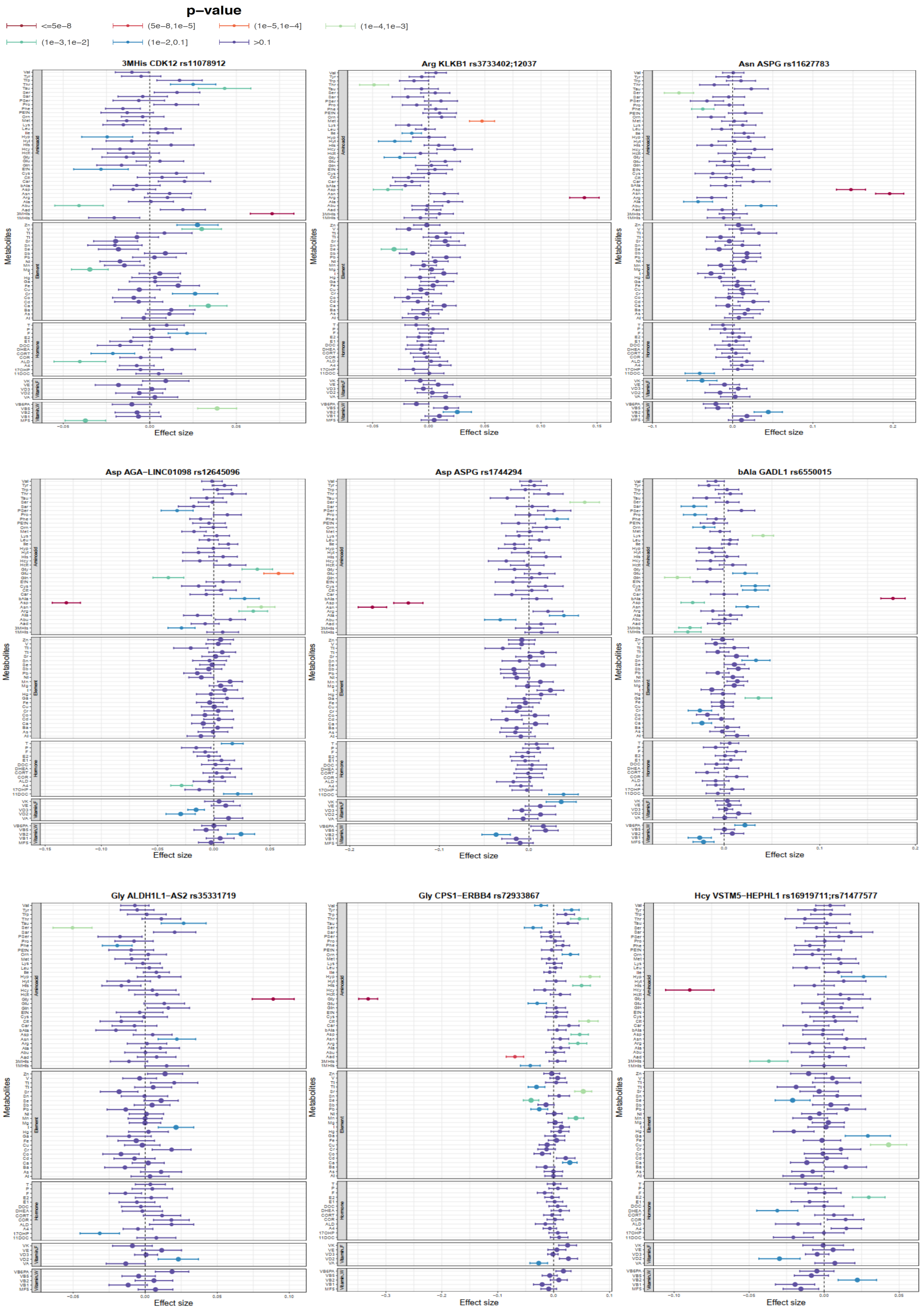
p−value
<=5e−8
(5e−8,1e−5]
(1e−5,1e−4]
(1e−4,1e−3]
(1e−3,1e−2]
(1e−2,0.1]
>0.1
3MHis CDK12 rs11078912
Arg KLKB1 rs3733402;12037
Asn ASPG rs11627783
Asp AGA−LINC01098 rs12645096
Asp ASPG rs1744294
bAla GADL1 rs6550015
Gly ALDH1L1−AS2 rs35331719
Gly CPS1−ERBB4 rs72933867
Hcy VSTM5−HEPHL1 rs16919711;rs71477577
Metabolites
Effect size

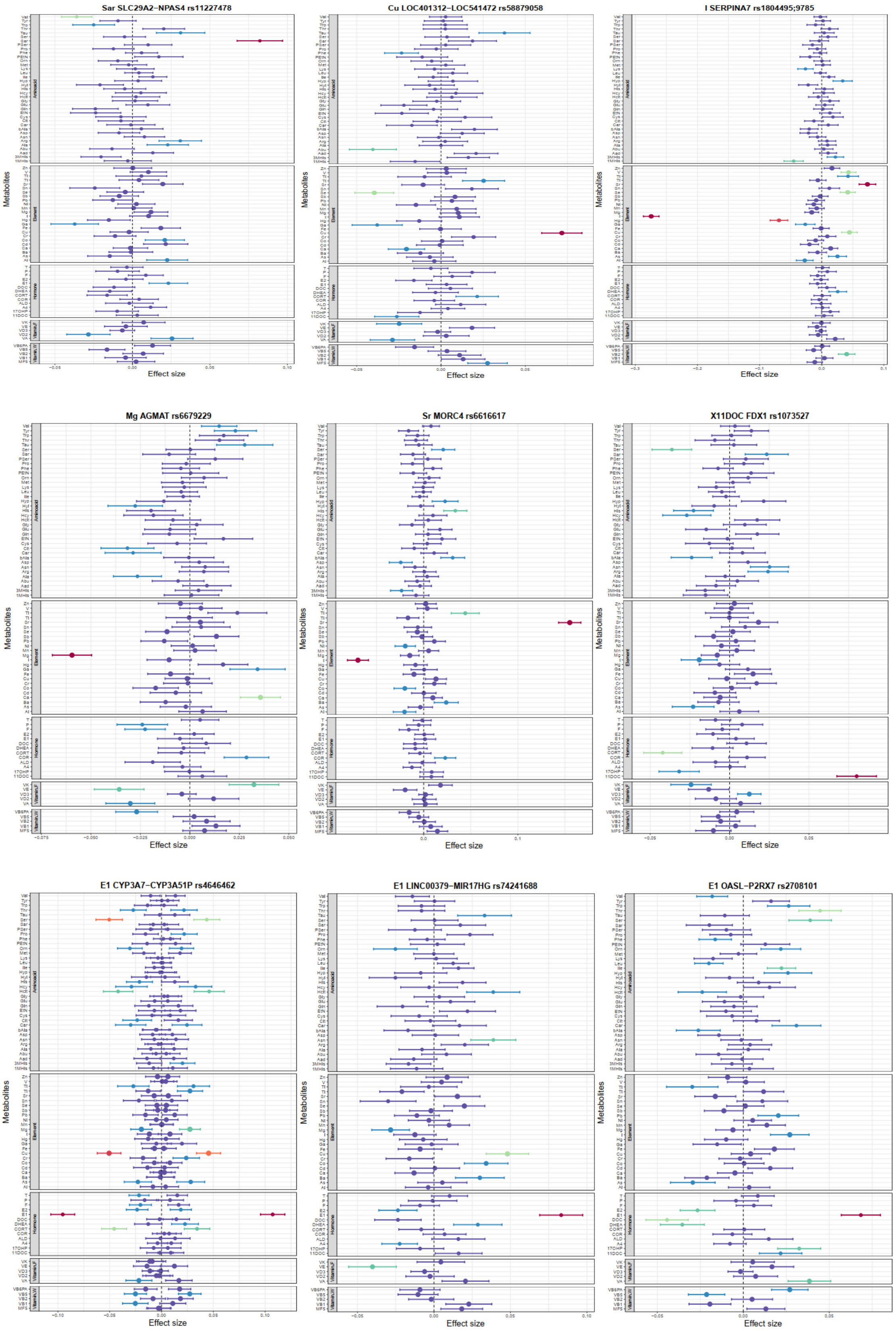
Sar SLC29A2−NPAS4 rs11227478
Cu LOC401312−LOC541472 rs58879058
I SERPINA7 rs1804495;9785
Mg AGMAT rs6679229
Sr MORC4 rs6616617
X11DOC FDX1 rs1073527
E1 CYP3A7−CYP3A51P rs4646462
E1 LINC00379−MIR17HG rs74241688
E1 OASL−P2RX7 rs2708101
Metabolites
Effect size
Aminoacid
Element
Hormone

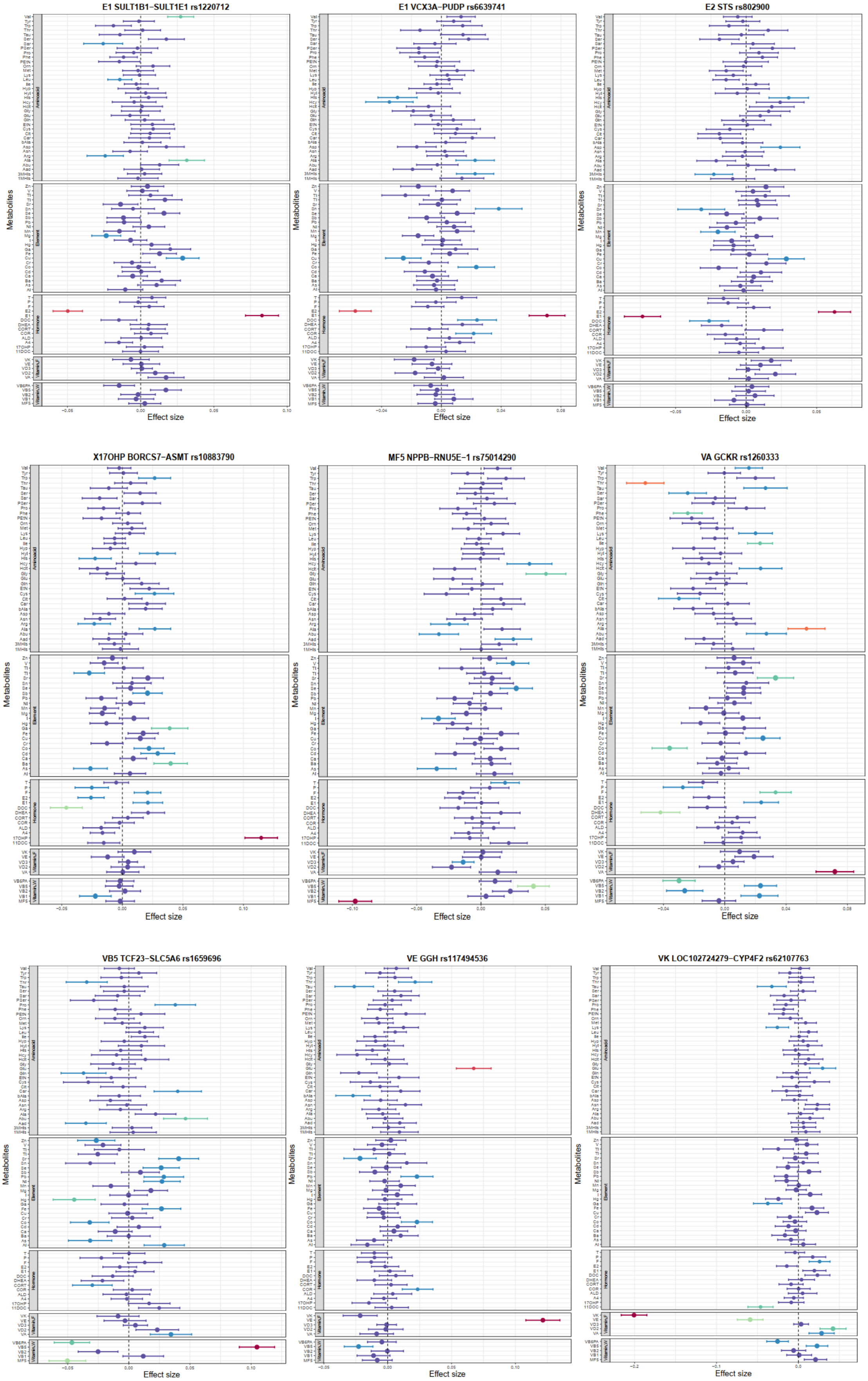
E1 SULT1B1−SULT1E1 rs1220712
E1 VCX3A−PUDP rs6639741
E2 STS rs802900
X17OHP BORCS7−ASMT rs10883790
MF5 NPPB−RNU5E−1 rs75014290
VA GCKR rs1260333
VB5 TCF23−SLC5A6 rs1659696
VE GGH rs117494536
VK LOC102724279−CYP4F2 rs62107763
Metabolites
Effect size
Aminoacid
Element
Hormone
Vitamin.F
Vitamin.W

**Figure S10. Forest plot for 27 loci affecting two or more metabolite molecules (p-value < 0.001)**

**a. (MM-Mg).(PP-UA).(PP-CR)**

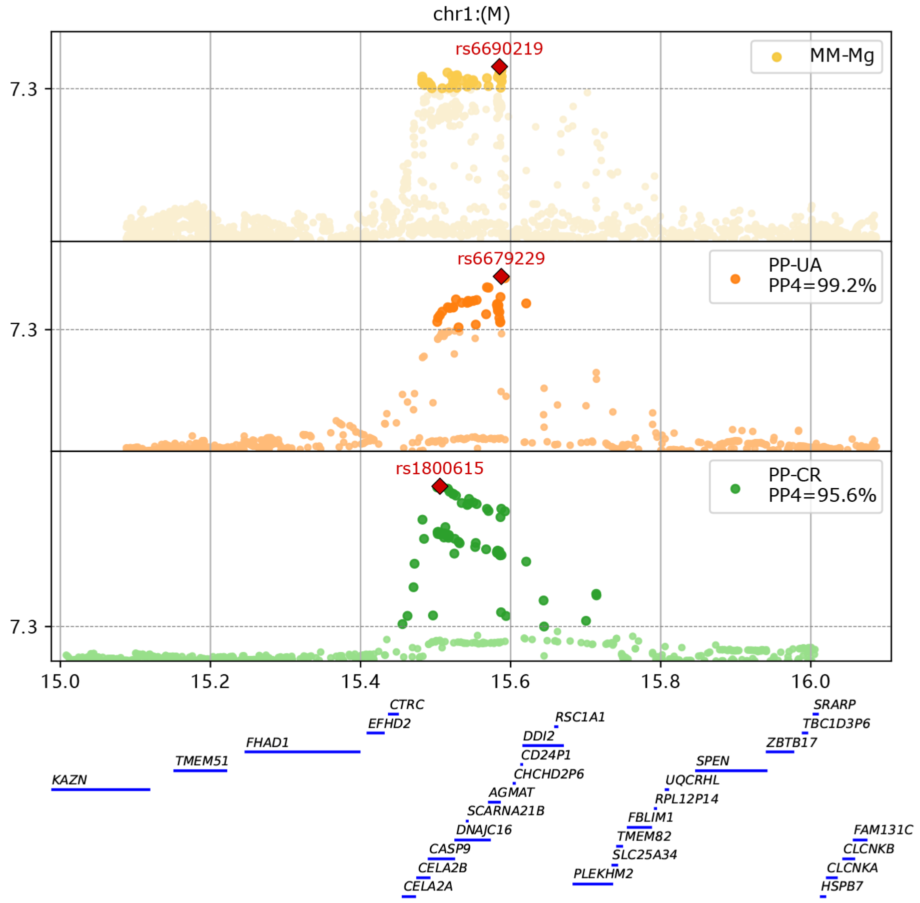

**b. (MM-VA). (PP-PA). (PP-CR)**

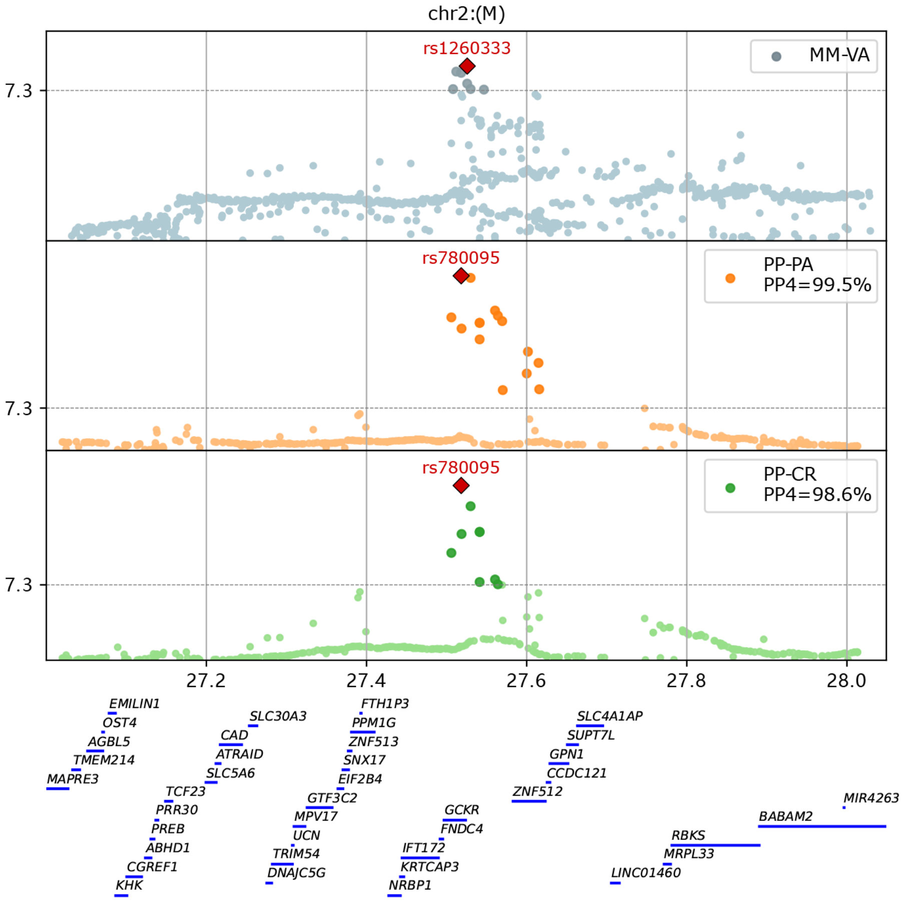

**c. (MM-E1).(MM-E2).(PP-MPV)**

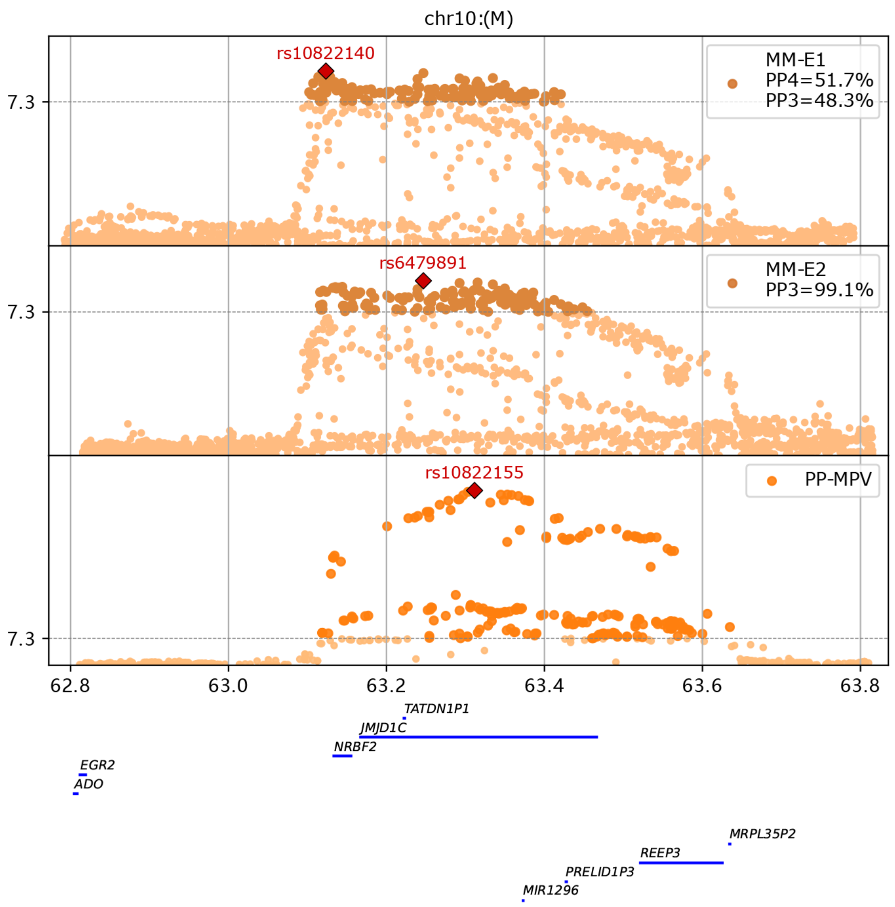

**d. (MM-3MHis). (PP-GLU_U)**

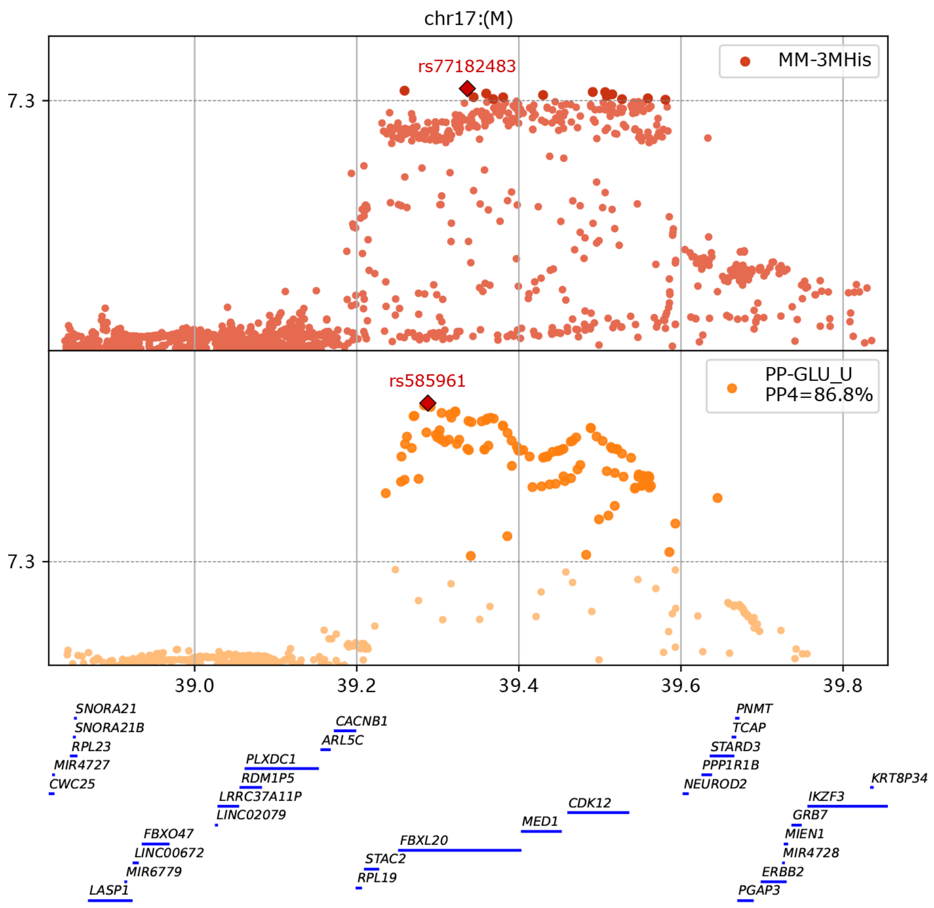

chr17:(M)
rs77182483
MM-3MHis
7.3
rs585961
PP-GLU_U
PP4=86.8%
7.3
39.0
39.2
39.4
39.6
39.8
SNORA21
SNORA21B
RPL23
MIR4727
CWC25
FBXO47
LINC00672
MIR6779
LASP1
CACNB1
ARL5C
PLXDC1
RDM1P5
LRRC37A11P
LINC02079
FBXL20
STAC2
RPL19
MED1
CDK12
PNMT
TCAP
STARD3
PPP1R1B
NEUROD2
KRT8P34
IKZF3
GRB7
MIEN1
MIR4728
ERBB2
PGAP3

**e. (MM-Fe).(PP-MCH)**

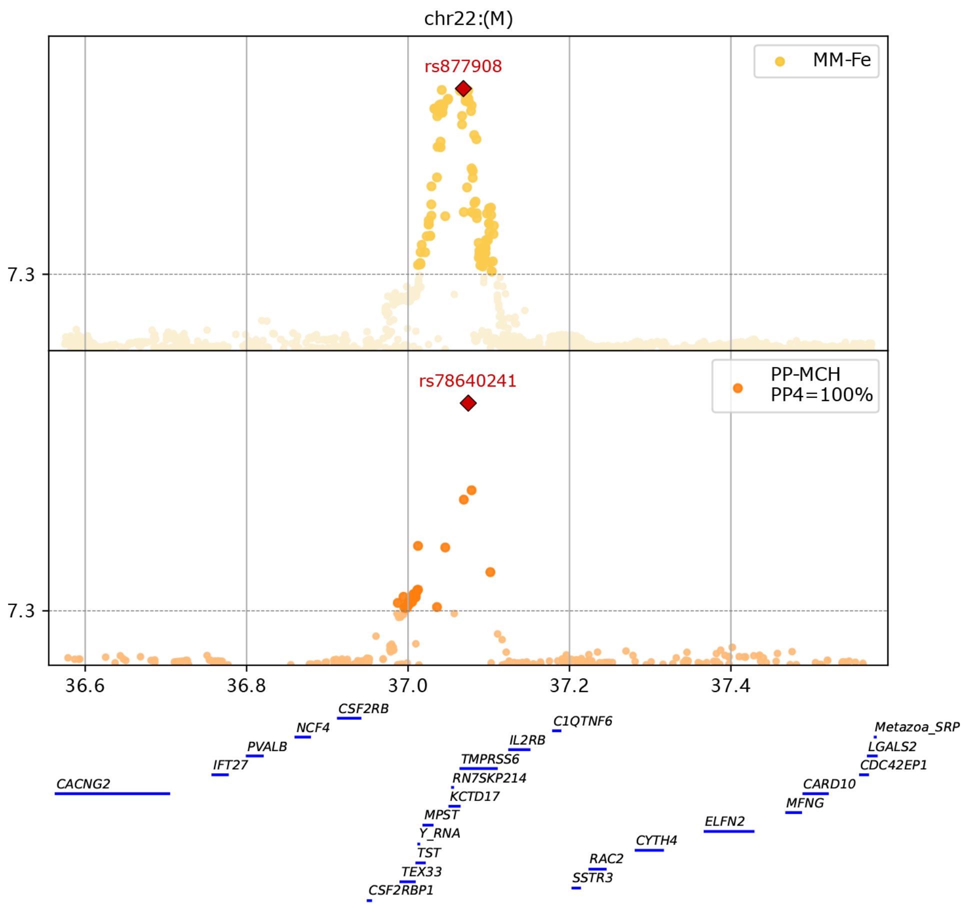

chr22:(M)
rs877908
MM-Fe
7.3
rs78640241
PP-MCH
PP4=100%
7.3
36.6
36.8
37.0
37.2
37.4
CSF2RB
NCF4
PVALB
IFT27
CACNG2
C1QTNF6
IL2RB
TMPRSS6
RN7SKP214
KCTD17
MPST
Y_RNA
TST
TEX33
CSF2RBP1
RAC2
SSTR3
CYTH4
ELFN2
MFNG
CARD10
CDC42EP1
LGALS2
Metazoa_SRP

**f. (MM-I).(PP-FT4)**

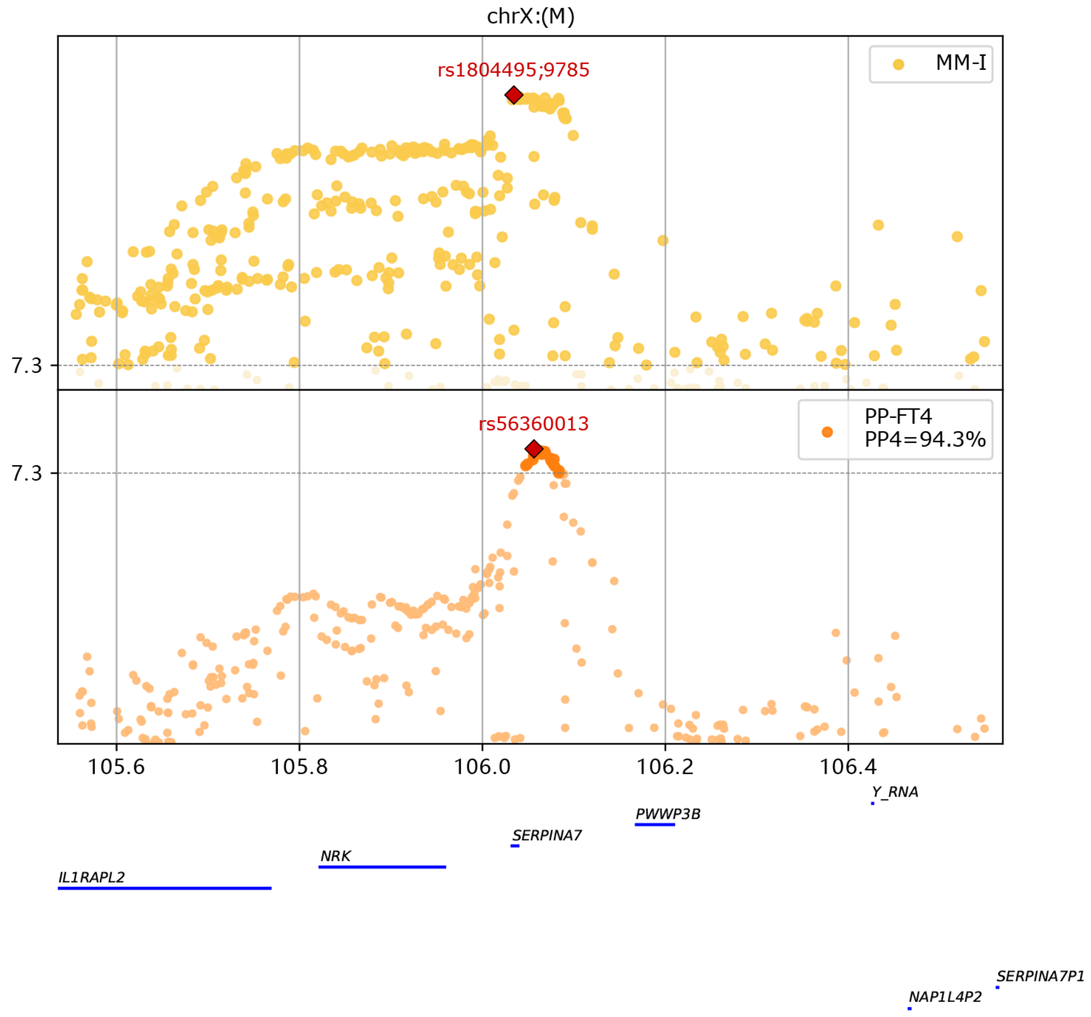

**Figure S11. Colocalization analysis reveals shared causal variants between seven maternal metabolites and pregnancy phenotypes.**

MM: maternal metabolites

PP: pregnancy phenotypes

Colocalization analysis was performed using the R coloc.abf function for the significant loci reported in this study. Pairs of phenotypes that shared posterior probabilities greater than 0.5 were considered as sharing a causal variant. P-values of multiple phenotypes were visualized with a gene annotation.
